# Genetic analyses of inflammatory polyneuropathy and chronic inflammatory demyelinating polyradiculoneuropathy identified candidate genes

**DOI:** 10.1101/2023.11.13.23298433

**Authors:** Zhaohui Du, Samuel Lessard, Tejaswi Iyyanki, Michael Chao, FinnGen, Timothy Hammond, Dimitry Ofengeim, Katherine Klinger, Emanuele de Rinaldis, Khader Shameer, Clément Chatelain

**Author notes:** **Corresponding author** Clément Chatelain, 350 Water Street, Cambridge, MA, USA 02141.

## Abstract

**Objective:** Chronic inflammatory demyelinating polyneuropathy (CIDP) is a rare, immune-mediated disorder in which an aberrant immune response causes demyelination and axonal damage of the peripheral nerves. Genetic contribution to CIDP is unclear and no genome-wide association study (GWAS) has been reported so far. In this study, we aimed to identify CIDP-related risk *loci*, genes and pathways.

**Methods:** To increase power, we first included all patients with a diagnosis of inflammatory polyneuropathy (IP) as cases. We performed a GWAS study using FinnGen R10 individual data and combined the results with GWAS from UK biobank (UKBB) using a fixed-effect meta-analysis. A total of 1,261 IP cases and 823,730 controls were included in the analysis. The second GWAS focused on CIDP patients and a total of 516 CIDP cases and 403,545 controls were included in the analysis. Stratified analyses by gender were also performed for both IP and CIDP. We performed gene-level analyses using transcriptome-wide mendelian randomization (TWMR) analysis, colocalization analysis, transcriptome-wide association study (TWAS) using S-PrediXcan and MAGMA to identify genes associated with IP and CIDP. Gene-set analyses were conducted using MAGMA to identify pathways that are related to IP and CIDP.

**Results:** In GWAS study, we identified one genome-wide significant loci at 20q13.33 for CIDP risk among women; the top variant located at the intron region of gene *CDH4*. TWMR, colocalization and S-PrediXcan analyses identified *DGKQ, GLDC, IDUA, SLAMF9* and *TMEM175* as candidate pathogenic genes for IP; genes *DIRAS1, GNG7*, and *SLC39A3* for CIDP; genes *DIRAS1, DCTN1*, and *ME1* for IP among males; and genes DIRAS1 and *ME1* for IP among women. MAGMA gene-set analyses identified a total of 18 pathways related to IP or CIDP.

**Conclusion:** Our study identified suggestive risk genes and pathways for IP and CIDP. Functional analysis should be conducted to further confirm these associations.

## Introduction

Chronic inflammatory demyelinating polyneuropathy (CIDP) is a rare, immune-mediated inflammatory polyneuropathy (IP) in which an aberrant immune response causes demyelination and axonal damage of the peripheral nerves [1]. CIDP is characterized by progressive weakness, sensory loss, and impaired reflexes in the limbs, which can lead to significant disability if left untreated. The incidence and prevalence of CIDP is 0.2-1.6 per 100,000 person-years and 0.8-10.3 per 100,000 persons, respectively, depending on geography and diagnostic criteria [2]. CIDP prevalence was higher among males and increased with age[3]. Despite the rarity of the disease, CIDP has a substantial impact on both physical and mental health[4], [5], which impairs patient quality of life[6]. CIDP is often practically viewed as the chronic counterpart of Guillain-Barré syndrome (GBS) due to various electrophysiological, histological, and immune similarities, despite differences in time course, mode of evolution, prognosis, and responsiveness to steroids.

While the etiology of CIDP is not fully understood, genetic factors are believed to play a role in disease susceptibility. Two early studies reported the occurrence of familial clustering of CIDP patients [7], [8]. A few candidate gene studies identified several genes associated with CIDP risk, such as the *GA13-16* homozygote genotype of the *SH2D2A* gene, which was found to be associated with increased CIDP risk (OR = 3.17) [9]. Additionally, several HLA polymorphisms, including the HLA-DRB1∗13 allele, have been found to be related to CIDP risk [10]. Gene expression studies also identified genes that are up-regulated in sural nerve biopsies from 8 patients of CIPD[11] and in skin biopsies from 11 CIDP patients[12]. These previous studies examined limited number of genes among very few patients. Given the current limited understanding of the genetic factors underlying CIDP susceptibility, a genome-wide association studies (GWAS) may provide valuable insights into the disease pathogenesis and facilitate the development of novel therapeutics.

In this study, we assembled data from two large biobanks and conducted the first genome- and transcriptome-wide association study for CIDP including a total of 516 cases and 403,545 controls. To identify genetic factors driving IP more generally and to potentially increase power, we also performed GWAS studies for IP, including a total of 1,261 IP patients and 823,730 controls. We also performed gender specified GWASs for both IP and CIDP.

## Results

### GWAS results

None of the GWAS analysis for IP or CIDP indicated evidence of inflation in association test statistics (e.g., due to confounding by population stratification; λ range 0.99 - 1.03, Supplementary figure 1). Genome-wide statistically significant associations were detected with 2 variants in one locus at 20q13.33 (Figure 1, Supplementary figure 2) in the CIDP GWAS analysis among women of FinnGen R10 participants. The lead variant, rs80339443 (OR = 4.37; 95% CI: 2.61, 7.33, P-value = 1.49 × 10^−8^), is located at the intron region of gene *CDH4*. The effect alle (C) of rs80339443 has a low frequency of 0.012 in the FinnGen population and is rare in other ancestral populations (EAF < 0.001 in Latino and African ancestry populations and = 0.01 in non-Finnish European population). No genome-wide signal was detected in any other GWAS analyses (Supplementary figures 2).

**Figure 1:**
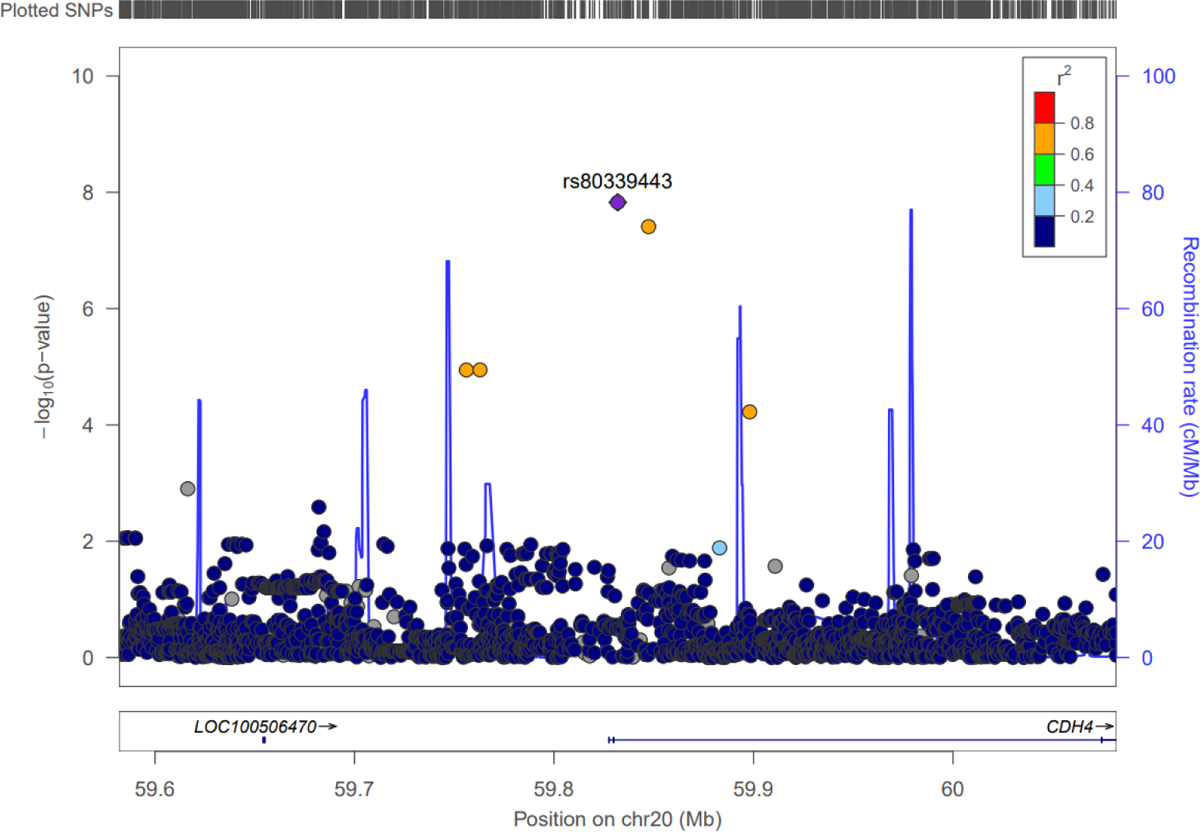
Regional association plots of the 20q13.33 risk regions for CIDP among women. Single-nucleotide polymorphisms (SNPs) are plotted by position (x-axis) and −log10 p-value (y-axis). r^2^ was estimated from EUR individuals in phase III 1000 Genomes Project (1KGP) data. The most statistically significant associated SNP (purple diamond) at 20q13.33 is rs80339443. The surrounding SNPs are colored to indicate pairwise correlation with the index SNP.

### MR and colocalization results

We performed MR analyses of IP using the meta-analysis GWAS summary statistics. 11 genes were identified as significantly associated genes at MR q-value < 0.05. We then conducted colocalization analyses to determine further the probability that SNPs associated with IP and eQTL shared causal genetic variants. IP GWAS signal colocalized with eQTLs of 5 genes in multiple tissues (probability >= 0.8), namely *DGKQ, IDUA, GLDC, SLAMF9* and *TMEM175* (Table 1, Supplementary table 1).

**Table 1.**
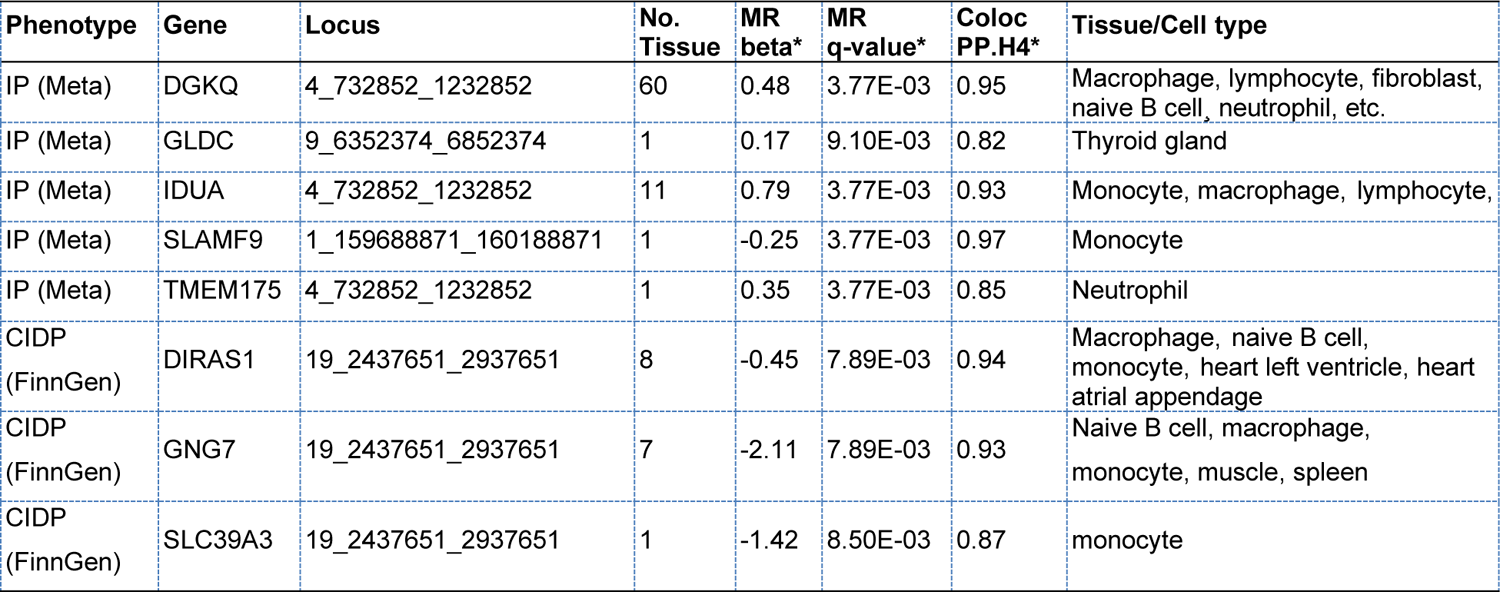
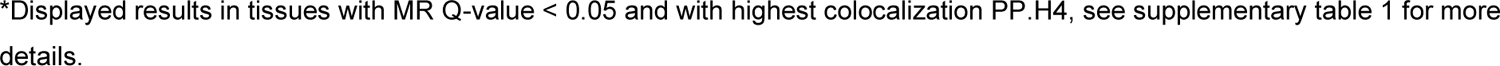
Genes that statistically significantly associated with inflammatory polyneuropathy (IP) and Chronic inflammatory demyelinating polyneuropathy (CIDP) in the MR (Q-value < 0.05) and Colocalization analyses (PP.H4 >= 0.8).

| Phenotype | Gene | Locus | No. Tissue | MR beta* | MR q-value* | Coloc PP.H4* | Tissue/Cell type |
| --- | --- | --- | --- | --- | --- | --- | --- |
| IP (Meta) | DGKQ | 4_732852_1232852 | 60 | 0.48 | 3.77E-03 | 0.95 | Macrophage, lymphocyte, fibroblast, naive B cell, neutrophil, etc. |
| IP (Meta) | GLDC | 9_6352374_6852374 | 1 | 0.17 | 9.10E-03 | 0.82 | Thyroid gland |
| IP (Meta) | IDUA | 4_732852_1232852 | 11 | 0.79 | 3.77E-03 | 0.93 | Monocyte, macrophage, lymphocyte, |
| IP (Meta) | SLAMF9 | 1_159688871_160188871 | 1 | -0.25 | 3.77E-03 | 0.97 | Monocyte |
| IP (Meta) | TMEM175 | 4_732852_1232852 | 1 | 0.35 | 3.77E-03 | 0.85 | Neutrophil |
| CIDP (FinnGen) | DIRAS1 | 19_2437651_2937651 | 8 | -0.45 | 7.89E-03 | 0.94 | Macrophage, naive B cell, monocyte, heart left ventricle, heart atrial appendage |
| CIDP (FinnGen) | GNG7 | 19_2437651_2937651 | 7 | -2.11 | 7.89E-03 | 0.93 | Naive B cell, macrophage, monocyte, muscle, spleen |
| CIDP (FinnGen) | SLC39A3 | 19_2437651_2937651 | 1 | -1.42 | 8.50E-03 | 0.87 | monocyte |

\*Displayed results in tissues with MR Q-value < 0.05 and with highest colocalization PP.H4, see supplementary table 1 for more details.

**Table 2.** Genes significantly associated with IP or CIDP risks in the S-PrediXcan analyses.

| Phenotype | Gene | Locus | Tissue | Z-score | P-value | FDR |
| --- | --- | --- | --- | --- | --- | --- |
| IP (Male) | DCTN1 | 2_73361154_75392087 | Brain Substantia nigra | 6.37 | 1.87E-10 | 3.59E-04 |
| IP (Male) | DIRAS1 | 19_1714567_3721372 | Esophagus Muscularis | 6.43 | 1.24E-10 | 3.59E-04 |
| IP (Female) | ME1 | 6_82210402_84431051 | EBV-transformed lymphocytes | 5.73 | 1.02E-08 | 7.82E-03 |
| IP (Female) | DIRAS1 | 19_1714567_3721372 | Esophagus Muscularis | 6.09 | 1.15E-09 | 1.47E-03 |
| IP (Female) | ME1 | 6_82210402_84431051 | Uterus | 5.99 | 2.11E-09 | 2.02E-03 |

Therefore, five potentially pathogenic genes with evidence of a shared genetic effect between the eQTL and IP risk were identified from MR and colocalization analyses. In the gender-stratified analysis, one gene was identified as significantly associated with IP risk among FinnGen women in the MR analysis but did not pass the threshold in the colocalization analysis.

In the MR analyses of CIDP using FinnGen GWAS summary statistics, 6 associated genes were identified with MR q-value < 0.05, among which 3 genes showed colocalization probability >= 0.8 in multiple tissues, including *DIRAS1, GNG7* and *SLC39A3* (Table 1, Supplementary table 1). In the gender-stratified analysis, one gene was significant in the MR analysis among FinnGen women but did not pass the threshold in the colocalization analysis. No significant gene was identified for either IP or CIDP in any other GWAS datasets.

### S-PrediXcan results

We also performed transcriptome-based analyses using tissue-specific gene-expression prediction models using S-PrediXcan to identify additional IP/CIDP associated genes. In total, we identified two additional genes associated with IP risk among FinnGen women, including *ME1* and *DIRASI*, and three genes associated with IP risk among FinnGen men, including *ME1*, *DCTN1* and *DIRAS1* at FDR q-value < 0.05.

### Gene and gene-set analyses results using MAGMA

We conducted gene- and pathway-based association analyses using MAGMA. No gene was identified in the gene level analyses for either IP or CIDP in either sex-combined or sex-stratified analyses. The gene-set analyses identified 3 significantly enriched pathways for sex-combined CIDP, and 3 and 2 pathways among females and males, respectively. For IP, a total of 10 pathways were identified in the GWAS analysis among women (Table 3).

**Table 3.**
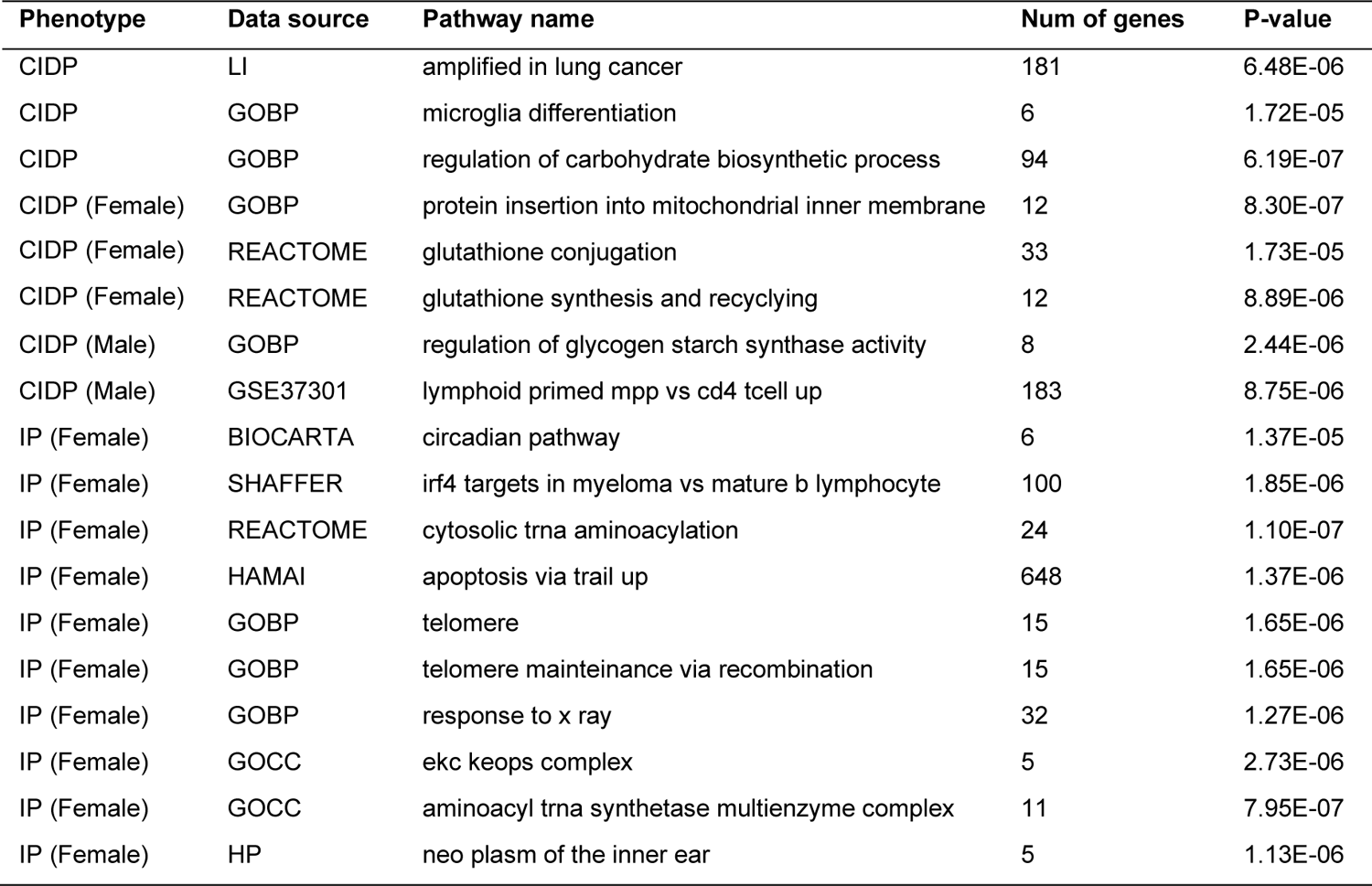
Pathways statistically significantly associated with IP or CIDP risks in MAGMA analysis.

### Target gene prioritization

After getting the list of candidate pathogenic genes associated with IP/CIDP risk using GWAS (nearest gene of GWAS *loci*), MR & colocalization, S-PrediXcan, and MAGMA gene-based analyses, we further gathered additional evidences that may support the pathogenicity of those genes through evaluating genetic associations with other potential related diseases (e.g. autoimmune diseases, neurological diseases and infections) using an in-house gene prioritization scheme, differential expressions in other diseases, bulk and single cell gene expression levels by tissues and cell types (Table 4, supplementary table 2-4, supplementary figure 3-5).

**Table 4:** Summary of genetic-/transcriptomic-associated phenotypes and highest expression tissues of candidate pathogenic genes.

| Gene | Phenotype | Discovery method | Phenotype with genetic association evidence (MR direction) | Phenotype with the most significant DEG (tissue: cell type) | Baseline gene expression | Highest expressed single cell, (percent of cells expressing the gene, %) |
| --- | --- | --- | --- | --- | --- | --- |
| CDH4 | CIDP (Female) | GWAS variant nearest gene | Cancer (+), Atopic dermatitis, COPD, Diabetic retinopathy, Nephritis | Alzheimer's disease (forebrain: pericyte cell) | Mainly in brain. | corticothalamic-projecting glutamatergic cortical neuron (59) |
| DGKQ | IP | MR + Coloc | Parkinson's disease (-) | Arthralgia (synovial tissue) | Ubiquitous expression, highest in brain - cerebellum. | corticothalamic-projecting glutamatergic cortical neuron (27) |
| GLDC | IP | MR + Coloc | Ashtma (+), Cardiomyopathy, Coxarthrosis:hip | Alzheimer's disease (forebrain: pericyte cell) | Mainly in liver. | trophectodermal cell (87) |
| IDUA | IP | MR + Coloc | Parkinson's disease (-) | frontotemporal lobar degeneration (skin: iPSC-derived neuron) | Ubiquitous expression, highest in brain. | endothelial progenitor cell (17) |
| SLAMF9 | IP | MR + Coloc |  | Asthma (peripheral blood: helper T cell) | Mainly restricted in hematopoietic tissues. | chondroblast (23) |
| TMEM175 | IP | MR + Coloc | Parkinson's disease (-), ALS (-) | Rheumatoid arthritis (synovial tissue) | Ubiquitous expression, highest in brain. | corticothalamic-projecting glutamatergic cortical neuron (70) |
| DIRAS1 | CIDP, IP (Male, Female) | MR + Coloc |  | Psoriasis vulgaris (skin: epidermal keratinocyte) | Highly expressed in brain and heart. | intratelencephalic-projecting glutamatergic cortical neuron (34) |
| GNG7 | CIDP | MR + Coloc | Gout | Clinically isolated syndrome (peripheral blood: PBMC) | Ubiquitous expression, predominantly in brain | plasmacytoid dendritic progenitor cell (74) |
| SLC39A3 | CIDP | MR + Coloc |  | Rheumatoid arthritis (synovial tissue) | Ubiquitous expression. | megakaryocyte progenitor cell (81) |
| DCTN1 | IP (Male) | S-PrediXcan |  | Celiac disease (intestinal mucosa) | Ubiquitous expression, predominantly in brain | fibrochondrocyte progenitor cell (72) |
| ME1 | IP (Male, Female) | S-PrediXcan | Myalgia and myositis | frontotemporal lobar degeneration (skin: iPSC-derived neuron) | Ubiquitous expression. | extratelencephalic-projecting glutamatergic cortical neuron (86) |

### CDH4

In genetic prioritization ranking with other potential related diseases, *CDH4* showed moderate evidence of correlation with cancer, with a significant positive MR (MR beta = 0.46, q-value < 0.05) association and high colocalization (maximum PP.H4 = 0.82) in CD8 naïve cell (Supplementary table 2). In differential gene expression analysis, *CDH4* is shown to be mostly upregulated in the breast tissue of breast cancer (log2-fold change = 2.32, adjusted P=1.1×10^-16^) and downregulated in the pericyte cell of forebrain tissue in Alzheimer’s disease (log2-fold change = −6.2, adjusted P=4.8×10^-52^) (Supplementary figure 3, Supplementary table 3). In bulk gene expression data, *CDH4* is mostly expressed in nervous system, with the highest expressed tissue being temporal lobe in brain tissue (Supplementary figure 4-5). In single cell data, *CDH4* is most expressed in corticothalamic-projecting glutamatergic cortical neuron, with 58.8% of cells expressing the gene, followed by intratelencephalic-projecting glutamatergic cortical neuron (41.3%) (Supplementary table 4).

### DGKQ

Gene *DGKQ* is associated with Parkinson’s disease (PD), which was supported by significant negative MR associations and high colocalization in four tissues, including fat, colon transverse, ovary, and liver. In differential gene expression analysis, *DGKQ* is shown to be mostly upregulated in the peripheral blood of congenital cytomegalovirus infection (log2-fold change = 1.27, adjust.P=5.8×10^-9^) and downregulated in the synovial tissue in arthralgia (log2-fold change = −3.3, adjust.P=1.4×10^-10^). In bulk gene expression data, gene *DGKQ* is mostly expressed in nervous system, with the highest expressed tissue being cerebellum in brain tissue. In single cell data, *DGKQ* is most expressed in corticothalamic-projecting glutamatergic cortical neuron, with 27% of cells expressing this gene.

### GLDC

Gene *GLDC* showed genetic evidence of association with Asthma (brain naïve tissue, MR beta = 1.0∼2.8, MR q-value < 0.05, max PP.H4 = 0.95) and Coxarthrosis (brain, MR beta = 0.56, MR q-value < 0.05, PP.H4 = 0.98; DLPFC naïve, MR beta = 0.71, MR q-value < 0.05, PP.H4 = 0.97). In differential gene expression analysis, *GLDC* is shown to be mostly upregulated in the pericyte cell of forebrain tissue of Alzheimer’s disease (log2-fold change = 5.82, adjust.P=8.8×10^-83^) and downregulated in iPSC-derived astrocyte cell of skin tissue in ALS (log2-fold change = −3.00, adjusted P=2.4×10^-16^). In bulk gene expression data, gene *GLDC* is expressed highest in liver, followed by EBV-Transformed Lymphocyte. In single cell data, *GLDC* is most expressed in trophectodermal cell, with 83% of cells expressing this gene.

### IDUA

Gene *IDUA* showed high genetic evidence relation with PD, with MR q-value < 0.05, high colocalization probability in 22 tissues (MR beta = −0.86 ∼ −0.30; highest PP.H4 = 0.99, in Nerve tibial and pancreas tissues) and coding variant association; and moderate genetic evidence with amyotrophic lateral sclerosis (ALS) (MR beta = −0.39 ∼ −0.11; PP.H4 >= 0.8 in 6 tissues, highest PP.H4 = 0.88, in muscle skeletal), arthrosis (MR beta = −0.01, pQTL MR) and dementia (MR beta = 0.05, pQTL MR). In differential gene expression analysis, *IDUA* is shown to be mostly upregulated in skeletal muscle of Dermatomyositis (log2-fold change = 1.77, adjusted P=1.6×10^-9^) and downregulated in the iPSC-derived neuron of skin tissue of frontotemporal lobar degeneration (log2-fold change = −1.36, adjusted P=3.8×10^-19^). In bulk gene expression data, gene *IDUA* is mostly expressed in nervous system, with the highest expressed tissue being cerebellum in brain tissue. In single cell data, *IDUA* is most expressed in endothelial progenitor cell, with 17% of cells expressing the gene. In addition, *IDUA* is an Informa target.

### TMEM175

Gene *TMEM175* showed strong genetic evidence of association with PD, with MR q-value < 0.05, moderate colocalization (MR beta = −0.22∼-0.12; max PP.H4 = 0.74 in neutrophil cell) and lead coding variant association; and moderate genetic evidence with ALS, with presence of a coding variant and being the lead ABC promoter contact.

Genes *TMEM175*, *IDUA* and *DGKQ* are at the same locus, and *TMEM175* is likely to be the causal gene driving PD at this locus based on the prioritization. In differential gene expression analysis, *TMEM175* is shown to be mostly upregulated in PBMC of peripheral blood in Parkinson’s disease (log2-fold change = 1.38, adjusted P=2.34×10^-6^) and downregulated in the synovial tissue of rheumatoid arthritis (log2-fold change = − 1.51, adjused P=5.7×10^-38^). In bulk gene expression data, gene *TMEM175* is mostly expressed in nervous system, with the highest expressed tissue being temporal Lobe in brain tissue. In single cell data, *TMEM175* is most expressed in intratelencephalic-projecting glutamatergic cortical neuron, with 70% of cells expressing the gene.

### GNG7

We did not identify genetic associations between *<u>GNG7</u>* and any disease using the genetic prioritization ranking scheme. In differential gene expression analysis, *GNG7* is shown to be mostly upregulated in iPSC-derived astrocyte of embryo tissue in Huntington’s disease (log2-fold change = 1.03, adjusted P=1.7×10^-3^) and downregulated in the PBMC cell of peripheral blood in clinically isolated syndrome (log2-fold change = − 2.53, adjusted P=2.1×10^-16^). In bulk gene expression data, gene *GNG7* is mostly expressed in nervous system, with the highest expressed tissue being putamen in brain tissue. In single cell data, *GNG7* is most expressed in plasmacytoid dendritic progenitor cell, with 74% of cells expressing the gene, followed by intratelencephalic-projecting glutamatergic cortical neuron (41.3%).

### DCTN1

Gene *DCTN1* did not show genetic evidence relation with any disease using the genetic prioritization ranking scheme. In differential gene expression analysis, *DCTN1* is shown to be mostly upregulated in PBMC of Clinically isolated syndrome (log2-fold change = 2.28, adjusted P=4.5×10^-11^) and downregulated in the intestinal mucosa in Celiac disease (log2-fold change = −1.52, adjusted P=3.3×10^-19^). In bulk gene expression data, gene *DCTN1* is mostly expressed in nervous system, with the highest expressed tissue being temporal lobe in brain tissue. In single cell data, *DCTN1* is most expressed in fibrochondrocyte progenitor cell, with 72% of cells expressing the gene.

### DIRAS1

Gene *DIRAS1* did not show genetic evidence for any disease using the genetic prioritization ranking scheme. In differential gene expression analysis, *DIRAS1* is shown to be mostly upregulated in the fibroblast cell of skin tissue of ataxia telangiectasia (log2-fold change = 1.28, adjusted P=1.9×10^-10^) and downregulated in epidermal keratinocyte cell of skin tissue in Psoriasis vulgaris (log2-fold change = −2.02, adjusted P=3.6×10^-20^). In bulk gene expression data, gene *DIRAS1* is mostly expressed in nervous system, with the highest expressed tissue being temporal lobe in brain tissue. In single cell data, *DIRAS1* is most expressed in intratelencephalic-projecting glutamatergic cortical neuron, with 34% of cells expressing the gene.

### ME1

Gene *ME1* did not show genetic evidence for any disease using the genetic prioritization ranking scheme. In differential gene expression analysis, *ME1* is shown to be mostly upregulated in the small airway epithelium tissue of COPD (log2-fold change = 1.98, adjusted P=1.8×10^-22^) and downregulated in the iPSC-derived neuron cell of skin tissue in frontotemporal lobar degeneration (log2-fold change = −3.95, adjusted P=8.7×10^-88^). In bulk gene expression data, gene *ME1* is mostly expressed in endocrine system, with the highest expressed tissue being adrenal Gland. In single cell data, *ME1* is most expressed in extratelencephalic-projecting glutamatergic cortical neuron, with 86% of cells expressing the gene.

### SLAMF9

Gene *SLAMF9* did not show genetic evidence for any disease using the genetic prioritization ranking scheme. In differential gene expression analysis, *SLAMF9* is shown to be mostly upregulated in the endothelial progenitor cell of peripheral blood in SLE (log2-fold change = 3.10, adjusted P=7.8×10^-6^) and downregulated in the iPSC-derived astrocyte cell of embryo tissue in Huntington’s disease (log2-fold change = − 4.14, adjusted P=1.1×10^-3^). In bulk gene expression data, gene *SLAMF9* is mostly expressed in Immune System and hematopoietic system, with the highest expressed tissue being alternatively activated macrophageIn single cell, *SLAMF9* is most expressed in chondroblast, with 23% of cells expressing the gene.

### SLC39A3

Gene *SLC39A3* did not show genetic evidence for any disease using the genetic prioritization ranking scheme. In differential gene expression analysis, *SLC39A3* is shown to be mostly upregulated in fibroblast cell of skin tissue in Ataxia telangiectasia (log2-fold change = 1.71, adjusted P=1.4×10^-13^) and downregulated in synovial tissue of rheumatoid arthritis (log2-fold change = −1.18, adjusted P=5.5×10^-25^). In bulk gene expression data, gene *SLC39A3* is mostly expressed in reproductive system, with the highest expressed tissue being testis tissue. In single cell data, *SLC39A3* is most expressed in megakaryocyte progenitor cell, with 81% of cells expressing the gene.

## Discussion

To our knowledge, this is the first GWAS studying CIDP genetic risk. We identified one novel genome-wide significant locus associated with CIDP risk among women in FinnGen. Leveraging gene-level TWMR, colocalization and S-PrediXcan analyses, we prioritized 5 novel candidate pathogenic genes for IP, and 3 for CIDP. In the sex-stratified analyses, 3 additional novel genes were identified for IP. We also identified a total of 18 pathways that are enriched in IP/CIDP genetic risk using gene-set analysis.

The genome-wide significant SNP rs80339443 located at the intron region of *CDH4* gene, which encodes the retinal-cadherin (R-cadherin) protein, a calcium-dependent cell adhesion protein. Cadherin has been indicated to play an important role in brain segmentation, neuronal outgrowth and muscle regeneration [13]–[18]. In addition, *CDH*4 is involved in cell-cell cohesion, inhibition of apoptosis and cell signaling, and tumorigenesis [19]. Some studies have suggested a potential role of *CDH*4 in neurodegeneration, related to axon genesis [20] and axon navigation and fasciculation [21].

Another prioritized gene identified in our analyses is *TMEM*175, which encodes a lysosomal proton channel protein that plays an important role in regulation of lysosomal pH and autophagy [22], as well as influence α-synuclein aggregation and mitochondrial function [23]. Autophagy plays a crucial role during neurodevelopment and neurogenesis, as well as at the synaptic zone [24]. Failure of autophagy in neurons can result in the accumulation of aggregate-prone proteins and neurodegeneration, and enhancing autophagy activity has been indicated as an efficient therapeutic strategy for neurodegenerative diseases such as Parkinson’s (PD) diseases [25]. Autophagy process has not been investigated in CIDP so far. Some animal models were studied on other polyneuropathy diseases. For example, one rat model of experimental autoimmune neuritis (EAN) suggested that autophagy activity is increased in nerve tissue of EAN rats, and the macroautophagy inhibitor 3-methyladenine, the inhibitor of autophagy, ameliorated the neurologic severity of EAN [26]. Another study using a rat model mimicking human CIDP, the chronic experimental autoimmune neuritis (chronic-EAN) model, demonstrate that prophylactic and therapeutic treatment of chronic-EAN rats with the CMA-targeting P140 peptide considerably ameliorates the clinical and biological course of the disease in chronic-EAN rats [27].

A major limitation in this study is the small sample size, with statistical power for genome-wide risk locus discovery being underpowered. In the exploratory genome-wide analyses, for alleles with MAF of 20%, there was only a priori adequate power (80%) to detect alleles with large effects (OR > 1.75 for CIDP and OR > 1.4 for IP) at genome-wide significance (P < 5 × 10^−8^). Nevertheless, it is challenging to obtain genetic data for a rare disease like CIDP and our study represents the first GWAS study for CIDP so far. More efforts are needed to pool more genetic data to achieve better statistical power.

In conclusion, our study discovered novel genetic risk *loci*, genes, and pathways that could provide further insights into biological mechanisms of IP and CIDP pathogenesis. Functional studies on these discoveries would be valuable to better understand CIDP pathophysiology and identify potential new treatments.

## Methods

### Study sample and GWAS

The study participants were from two independent large-scale biobanks: the UK Biobank (UKBB) and FinnGen. For UKBB, we obtained publicly available GWAS summary statistics of IP from Pan-UK Biobank (https://pan.ukbb.broadinstitute.org/, 2017 release). This GWAS was performed in 346 IP cases with ICD-10 diagnosis of G61 and 420,185 healthy controls. Per-allele odds ratios (ORs) and P-value of each variant were estimated using unconditional logistic regression, adjusting for age, sex, age * sex, age^2^, age^2^ * sex, and the first 10 principal components (PC). In the FinnGen Consortium (release 10), we defined IP cases as individuals with G61[0,1,8,9] (ICD - 10), 357[0,7,8,9] (ICD-9) or 354[01,08,09] (ICD-8) diagnoses, which included 915 patients, and CIDP cases as individuals with G61[8–9] (ICD-10), 357[8,9] (ICD-9) or 354[08,09](ICD-8) diagnoses, which included 516 patients. The controls for both IP and CIDP were defined as individuals without any G60-G64 (icd-10), 356-357 (icd-9) or 354 (icd-8) diagnoses, which included 403,545 participants. The analyses were adjusted for age, sex, the first 10 PCs, and genotyping batch and were performed using the REGENIE pipeline in FinnGen sandbox [28].

Fixed effect meta-analyses with inverse variance weights were performed using the METAL software [29] to combine GWAS results from UKBB and FinnGen. The meta-analyses were performed both for the sex-combined and sex-specific GWAS.

### Post-GWAS analyses Variant annotation

We annotated the impact of all variants with P<1×10^-6^ using the variant effect predictor (VEP v102)[30] with the following options: --everything --offline --check_existing. Coding variants were defined as those impacting protein coding transcript annotated as missense variant or predicted to have “high” impact. We also retrieved predicted gain or loss of function variants6. We also mapped variants to genes using activity-by-contact (ABC) enhancer-promoter interactions[31]. We used “ABCmax” to represent variant-gene pairs with the highest ABC score.

### Mendelian randomization and colocalization

We conducted transcriptome wide mendelian randomization (TWMR) using the TwoSampleMR R package [32]. The following exposures were considered: protein quantitative trait loci (pQTL) from Sun et al [33], and expression quantitative trait loci from Blueprint [34], eQTLGen [35] and other datasets from the EBI eQTL catalogue [34]–[45]. For each of those studies, variants with a MAF < 1% were excluded. Variants were clumped using PLINK2 87 using the options –clump-p1 1 –clump-p2 1 –clump-r2 0.01 – clump-kb 10000. Only genes 250kb around significant loci were considered in this analysis. For each QTL, independent variants with P < 1×10^-4^ were used as instruments. When more than one instrument was present, the inverse variant weighted approach was used, otherwise the Wald Ratio approach was used. False Discovery Rate (FDR) corrections were applied to adjust P-values in multiple testing, and a threshold of FDR < 0.05 was used to determine significance.

For significant TWMR results, we performed colocalization analyses to detect shared causal SNPs between molecular-QTL and IP/CIDP risk using the coloc R package with default priors [46]. We used a posterior probability threshold of H4 (PP.H4) >= 0.8 for colocalization, and genes with both significant MR and colocalization were considered as potential targeted molecular.

### Gene-Based Association Using Transcriptomic Data With S-PrediXcan

We used S-PrediXcan [47] to integrate cis-eQTL information with GWAS summary statistics to identify genes whose genetically predicted expression levels are associated with IP/CIDP risk. We employed the multivariate adaptive shrinkage (MASHR) models, which were precomputed from 49 tissues in the Gene-Tissue Expression Project (GTEx) version 8 [15], as our tissue-specific prediction models. These models were obtained from http://predictdb.org/ and were trained in tissues from European-ancestry population.

### Gene and gene-set analyses using MAGMA

We performed gene and gene-set level analyses using MAGMA (v1.10) [48]. SNP annotation to genes was done using the 1000 Genomes Phase 3 European panel and GRCh38 (downloaded from http://ctglab.nl/software/magma). Gene sets were downloaded from the Molecular Signatures Database (MSigDB) [49], [50], including 7763 gene sets in Gene Ontology (GO), 186 in Kyoto Encyclopedia of Genes and Genomes (KEGG), 292 in BioCarta, 1,635 in Reactome and 4,872 in ImmuneSigDB.

### Associations of identified genes with other phenotypes

For IP/CIDP related genes identified from GWAS (nearest gene in the GWAS *loci*), MR & Colocalization, MAGMA, and S-PrediXcan analyses, we then searched for other autoimmune or neurological diseases that showed evidence of association with the focal genes. We also included some infection diseases (e.g. HIV, Hepatitis B or C) because CIDP may be triggered by an infection in some case report, and cancers because some studies reported an increased incidence of cancer in patients with CIDP. We employed an in-house gene prioritization scheme, which contained precalculated results across 6,918 GWAS from 4 sources, including UK Biobank, FinnGen release 10 (R10), EBI GWAS catalog, and meta-analyses of UK Biobank, FinnGen R10, and Estonian biobank. This gene-ranking scheme used a combinatory approach to prioritize disease putative genes based on MR, eQTL colocalization, activity-by-contact (ABC) enhancer-promoter interactions, and variant annotations. This approach mitigates the pleiotropic effect of eQTL while retaining important information about directionality and has been shown to be enriched for “gold standard” genes and drug targets with successful clinical trial.

### Differential expression in other diseases

We further examined whether the identified IP/CIDP related genes are differentially expressed in diseased compared to normal tissues of other autoimmune, neurological diseases, infection diseases and cancer using bulk transcriptomics data. Differential gene expressions (DGE) were extracted from integrated transcriptomics service from Qiagen Omicsoft database (QIAGEN OmicSoft DiseaseLand[51]). We extracted both microarray and RNA-seq based transcriptomic (TxP) studies and disease comparisons/contrasts available for all studies in the DiseaseLand in the immunology and inflammation, neurodegenerative and metabolic diseases[52]. Common ontologies were applied to all samples, tissues, diseases, treatments, and comparison types to systematize database curation. Database raw data was derived from published repositories including gene expression omnibus (GEO) and ArrayExpress (https://www.ebi.ac.uk/biostudies/arrayexpress). In total, the integrated database consists of >250,000 samples with TxP profiles, 32,000 study comparison contrasts in ∼ 700 disease areas. The contrasts that were of interest to us are “Disease.vs.Normal” which we subsequently used to perform meta-analysis.

Several studies have delved into the exploration of integrated transcriptomics data through various meta-analyses [27], [28]. We opted for the Fisher’s exact test for its straightforward implementation, heightened sensitivity to small sample sizes, non-parametric nature, and platform independence across transcriptomics techniques (such as microarray and RNA-seq). Overall, meta-analysis examined whether a gene_i is a recurrent DGE enriched for a given disease, tissue, or sample (e.g.: Alzheimer’s disease and brain tissue) when compared to recurrency of the gene in the entire database. We constructed a contingency table as outlined in the methodology suggested earlier by Wang et al.[53] where we used a cut-off of adjusted P values (P-adj<0.05) & abs(log2Fold-change)>1.0 to define a DGE, and at least 70% of studies having the same direction of log2Fold-Change (positive or negative). A recurrent DGE enriched across different studies increases the confidence of the gene in playing a role in the disease mechanism of action (MoA).

### Gene expression levels by cell types and tissues

We then assessed the expression levels of the identified IP/CIDP related genes by cell types and tissues. Bulk gene expression data were retrieved from GTEx and single cell data were obtained from the BioTuring Single-cell gene expression database [29].

## ETHICS APPROVAL AND CONSENT TO PARTICIPATE

Patients and control subjects in FinnGen provided informed consent for biobank research, based on the Finnish Biobank Act. Alternatively, separate research cohorts, collected prior the Finnish Biobank Act came into effect (in September 2013) and start of FinnGen (August 2017), were collected based on study-specific consents and later transferred to the Finnish biobanks after approval by Fimea, the National Supervisory Authority for Welfare and Health. Recruitment protocols followed the biobank protocols approved by Fimea. The Coordinating Ethics Committee of the Hospital District of Helsinki and Uusimaa (HUS) approved the FinnGen study protocol Nr HUS/990/2017.

The FinnGen study is approved by Finnish Institute for Health and Welfare (permit numbers: THL/2031/6.02.00/2017, THL/1101/5.05.00/2017, THL/341/6.02.00/2018, THL/2222/6.02.00/2018, THL/283/6.02.00/2019, THL/1721/5.05.00/2019, THL/1524/5.05.00/2020, and THL/2364/14.02/2020), Digital and population data service agency (permit numbers: VRK43431/2017-3, VRK/6909/2018-3, VRK/4415/2019-3), the Social Insurance Institution (permit numbers: KELA 58/522/2017, KELA 131/522/2018, KELA 70/522/2019, KELA 98/522/2019, KELA 138/522/2019, KELA 2/522/2020, KELA16/522/2020 and Statistics Finland (permit numbers: TK-53-1041-17 and TK-53-90-20).

The Biobank Access Decisions for FinnGen samples and data utilized in FinnGen Data Freeze 6 include: THL Biobank BB2017_55, BB2017_111, BB2018_19, BB_2018_34, BB_2018_67, BB2018_71, BB2019_7, BB2019_8, BB2019_26, BB2020_1, Finnish Red Cross Blood Service Biobank 7.12.2017, Helsinki Biobank HUS/359/2017, Auria Biobank AB17-5154, Biobank Borealis of Northern Finland_2017_1013, Biobank of Eastern Finland 1186/2018, Finnish Clinical Biobank Tampere MH0004, Central Finland Biobank 1-2017, and Terveystalo Biobank STB 2018001.

UK Biobank has received ethical approval from the NHS National Research Ethics Service North West (approval numbers 11/NW/0382 and 16/NW/0274). All participants provided written informed consent.

## Data Availability

The UK Biobank GWAS is available through https://pan.ukbb.broadinstitute.org/. The FinnGen GWAS are available through https://www.finngen.fi/en/access_results.

## UK Biobank

This research has been conducted using the UK Biobank Resource (www.ukbiobank.ac.uk), a large-scale biomedical database and research resource containing genetic, lifestyle and health information from 500,000 UK participants. UK Biobank is supported by its founding funders the Wellcome Trust and UK Medical Research Council, as well as the Department of Health, Scottish Government, the Northwest Regional Development Agency, British Heart Foundation and Cancer Research UK. The UK biobank pan-ancestry analysis was conducted under project ID 31063 (https://pan.ukbb.broadinstitute.org).

## FinnGen

We want to acknowledge the participants and investigators of FinnGen study. The FinnGen project is funded by two grants from Business Finland (HUS 4685/31/2016 and UH 4386/31/2016) and the following industry partners: AbbVie Inc., AstraZeneca UK Ltd, Biogen MA Inc., Bristol Myers Squibb (and Celgene Corporation & Celgene International II Sàrl), Genentech Inc., Merck Sharp & Dohme LCC, Pfizer Inc., GlaxoSmithKline Intellectual Property Development Ltd., Sanofi US Services Inc., Maze Therapeutics Inc., Janssen Biotech Inc, Novartis Pharma AG, and Boehringer Ingelheim International GmbH. Following biobanks are acknowledged for delivering biobank samples to FinnGen: Auria Biobank (www.auria.fi/biopankki), THL Biobank (www.thl.fi/biobank), Helsinki Biobank (www.helsinginbiopankki.fi), Biobank Borealis of Northern Finland (https://www.ppshp.fi/Tutkimus-ja-opetus/Biopankki/Pages/Biobank-Borealis-briefly-in-English.aspx), Finnish Clinical Biobank Tampere (www.tays.fi/en-US/Research_and_development/Finnish_Clinical_Biobank_Tampere), Biobank of Eastern Finland (www.ita-suomenbiopankki.fi/en), Central Finland Biobank (www.ksshp.fi/fi-FI/Potilaalle/Biopankki), Finnish Red Cross Blood Service Biobank (www.veripalvelu.fi/verenluovutus/biopankkitoiminta), Terveystalo Biobank (www.terveystalo.com/fi/Yritystietoa/Terveystalo-Biopankki/Biopankki/) and Arctic Biobank (https://www.oulu.fi/en/university/faculties-and-units/faculty-medicine/northern-finland-birth-cohorts-and-arctic-biobank). All Finnish Biobanks are members of BBMRI.fi infrastructure (www.bbmri.fi). Finnish Biobank Cooperative - FINBB (https://finbb.fi/) is the coordinator of BBMRI-ERIC operations in Finland. The Finnish biobank data can be accessed through the Fingenious® services (https://site.fingenious.fi/en/) managed by FINBB.

### Estonian Biobank

Estonian Biobank research was supported by the European Union through Horizon 2020 research and innovation programme under grant no 810645 and through the European Regional Development Fund project no. MOBEC008, by the Estonian Research Council grant PUT (PRG1291, PRG687 and PRG184) and by the European Union through the European Regional Development Fund project no. MOBERA21 (ERA-CVD project DETECT ARRHYTHMIAS, GA no JTC2018-009), Project No. 2014-2020.4.01.15-0012 and Project No. 2014-2020.4.01.16-0125.

