## Supplementary figure 1-2 for "Genetic analyses of inflammatory polyneuropathy and chronic inflammatory demyelinating polyradiculoneuropathy identified candidate genes"

a.1

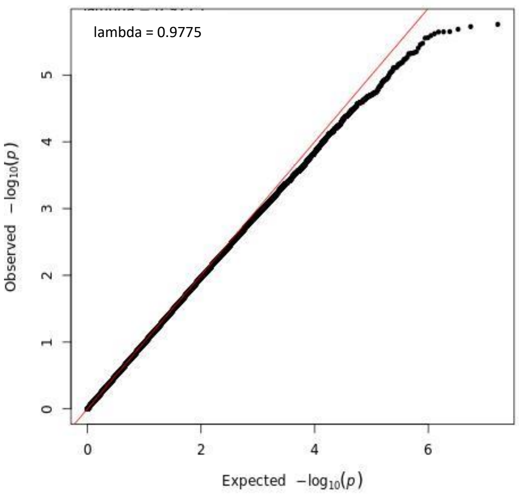
lambda = 0.9775
Observed $-\log_{10}(p)$
Expected $-\log_{10}(p)$

a.2

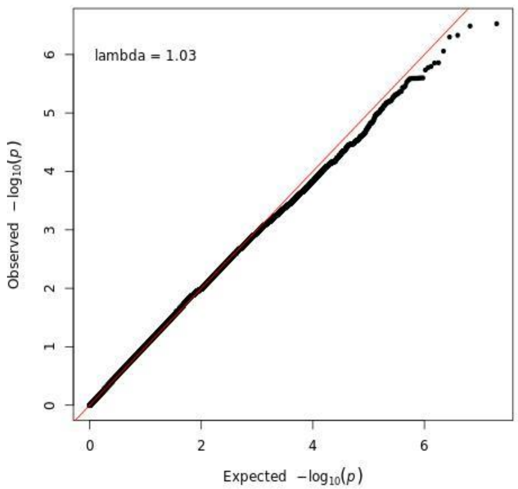
lambda = 1.03
Observed $-\log_{10}(p)$
Expected $-\log_{10}(p)$

a.3

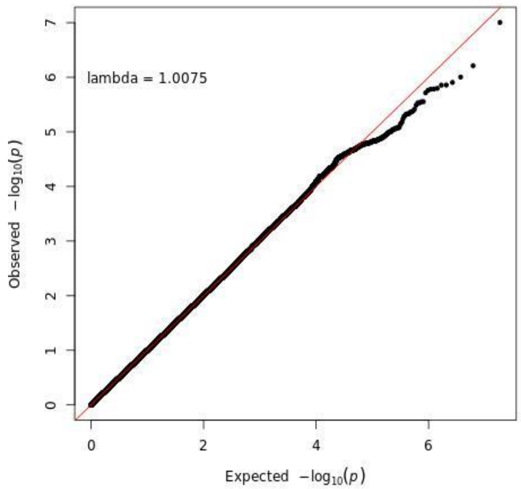
lambda = 1.0075
Observed $-\log_{10}(p)$
Expected $-\log_{10}(p)$

b.1

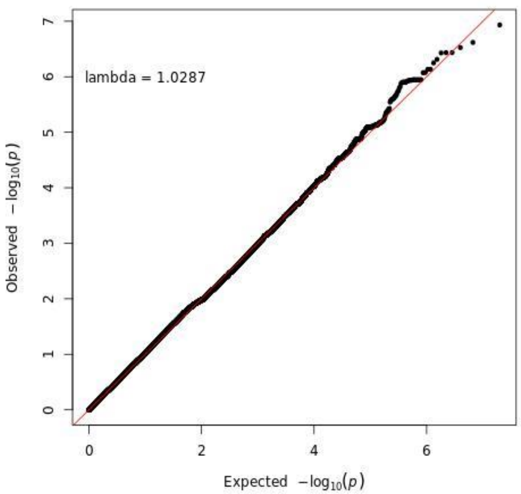
lambda = 1.0287
Observed $-\log_{10}(p)$
Expected $-\log_{10}(p)$

b.2

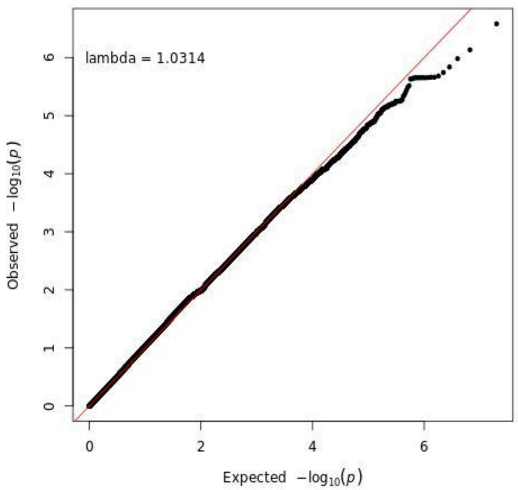
lambda = 1.0314
Observed $-\log_{10}(p)$
Expected $-\log_{10}(p)$

c.1

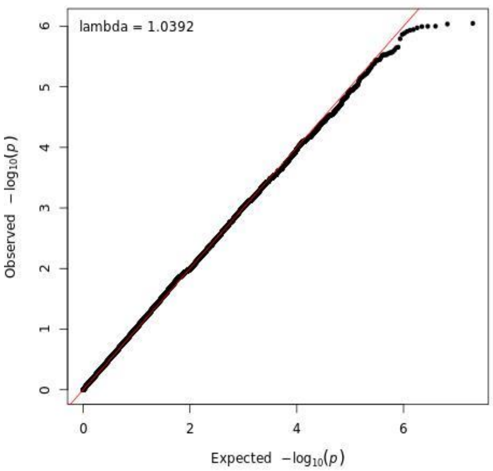


c.2

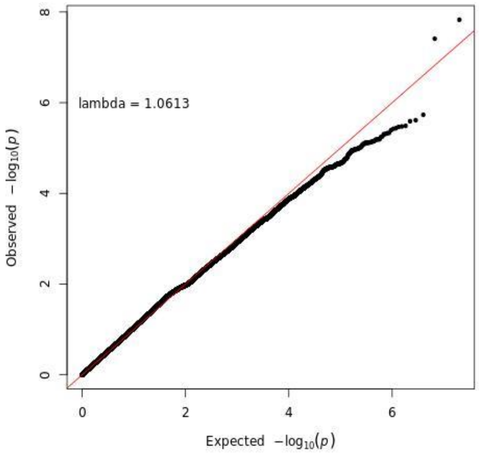


c.3

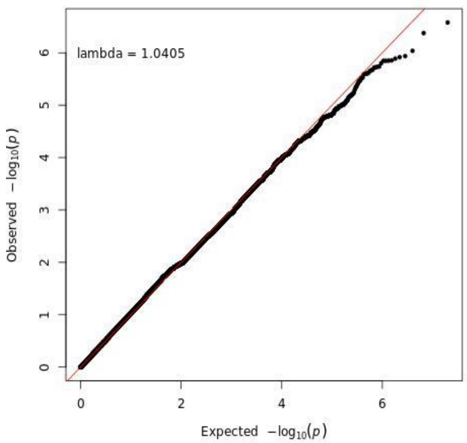


**Supplementary figure 1**. QQ plot of GWAS analyses for inflammatory polyneuropathy (IP) (a.1 Meta-analysis of ) and Chronic inflammatory demyelinating polyneuropathy (CIDP). QQ plot shows all genotyped and imputed results, excluding SNPs with MAF < 0.01 and imputation score <0.8. x-Axis is expected −log10(p-value) from normal distribution, y-axis is −log10(p-value) from our GWAS analyses. a.1. Meta-analysis of IP in UK Biobank and FinnGen R10; a.2. GWAS in IP of FinnGen R10; a.3. GWAS of IP in UK Biobank. b.1. GWAS of IP in females of FinnGen R10; b.2 GWAS of IP in males of FinnGen R10. C.1. GWAS of CIDP in FinnGen R10; c.2 GWAS of CIDP in females of FinnGen R10; c.3 GWAS of CIDP in males of FinnGen R10.

a.1

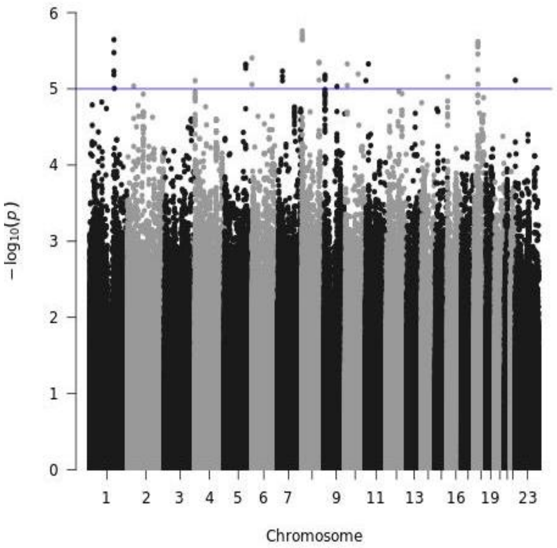
-log10(p)
Chromosome

a.2

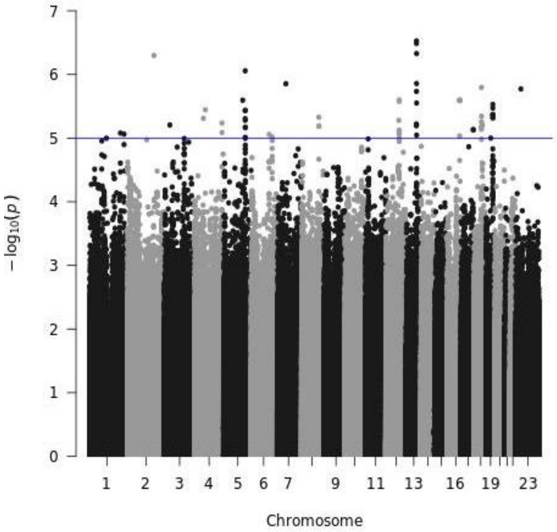
-log10(p)
Chromosome

a.3

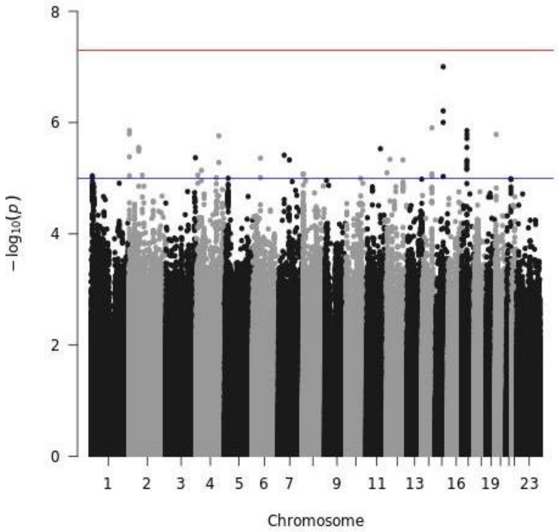
-log10(p)
Chromosome

b.1

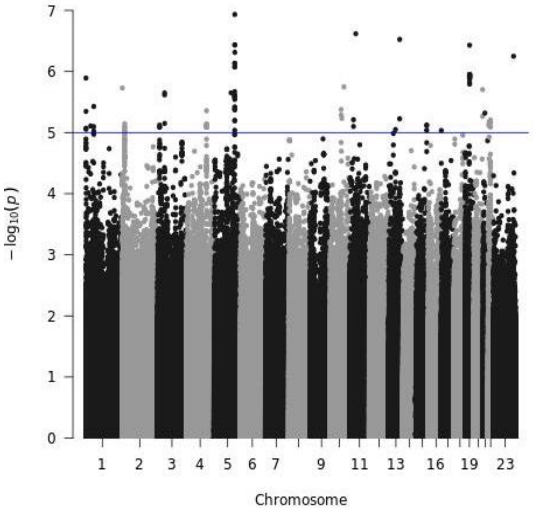
-log10(p)
Chromosome

b.2

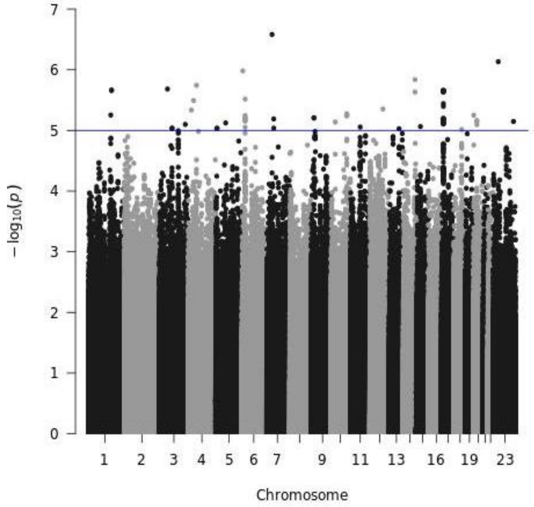
-log10(p)
Chromosome

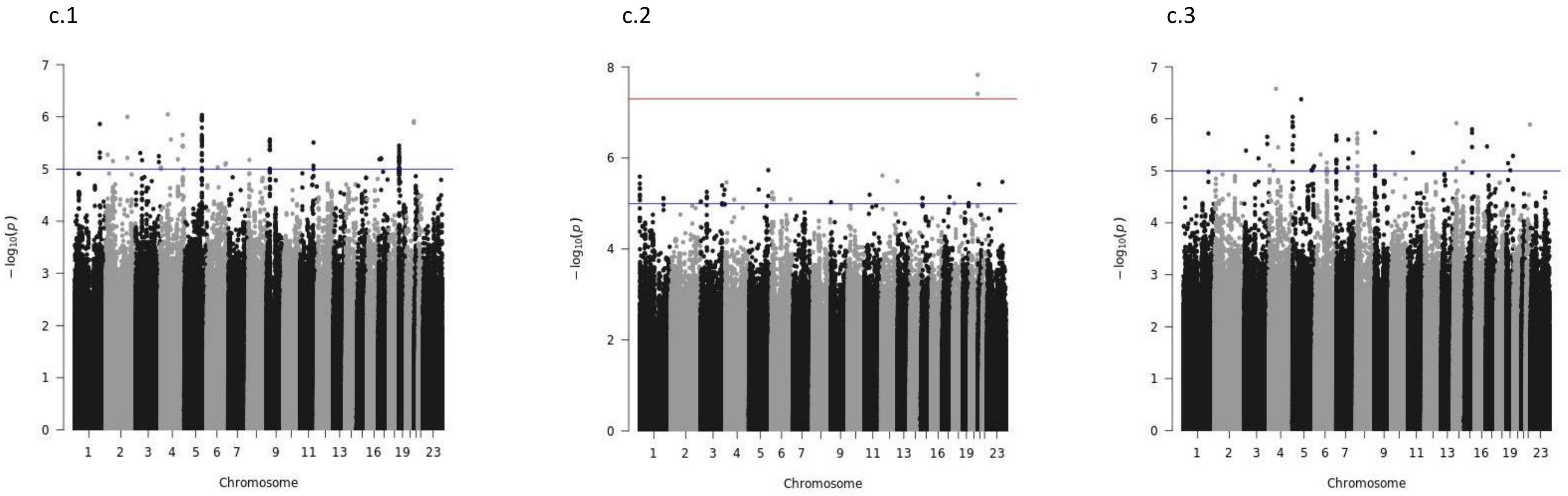


**Supplementary figure 2**. Manhattan plot of GWAS for inflammatory polyneuropathy (IP) and Chronic inflammatory demyelinating polyneuropathy (CIDP) in males. Manhattan plot shows all genotyped and imputed results, excluding SNPs with MAF < 0.01 and imputation score < 0.8. x-Axis is chromosome position, y-axis is −log10(p-value). The red line represents the genome-wide significant cut-off value of $p = 5 \times 10^{-8}$, and blue line represents suggestive significance level at $1 \times 10^{-5}$. a.1. Meta-analysis of IP in UK Biobank and FinnGen R10; a.2. GWAS in IP of FinnGen R10; a.3. GWAS of IP in UK Biobank. b.1. GWAS of IP in females of FinnGen R10; b.2 GWAS of IP in males of FinnGen R10. C.1. GWAS of CIDP in FinnGen R10; c.2 GWAS of CIDP in females of FinnGen R10; c.3 GWAS of CIDP in males of FinnGen R10.
