## Supplementary figure 3 for "Genetic analyses of inflammatory polyneuropathy and chronic inflammatory demyelinating polyradiculoneuropathy identified candidate genes"

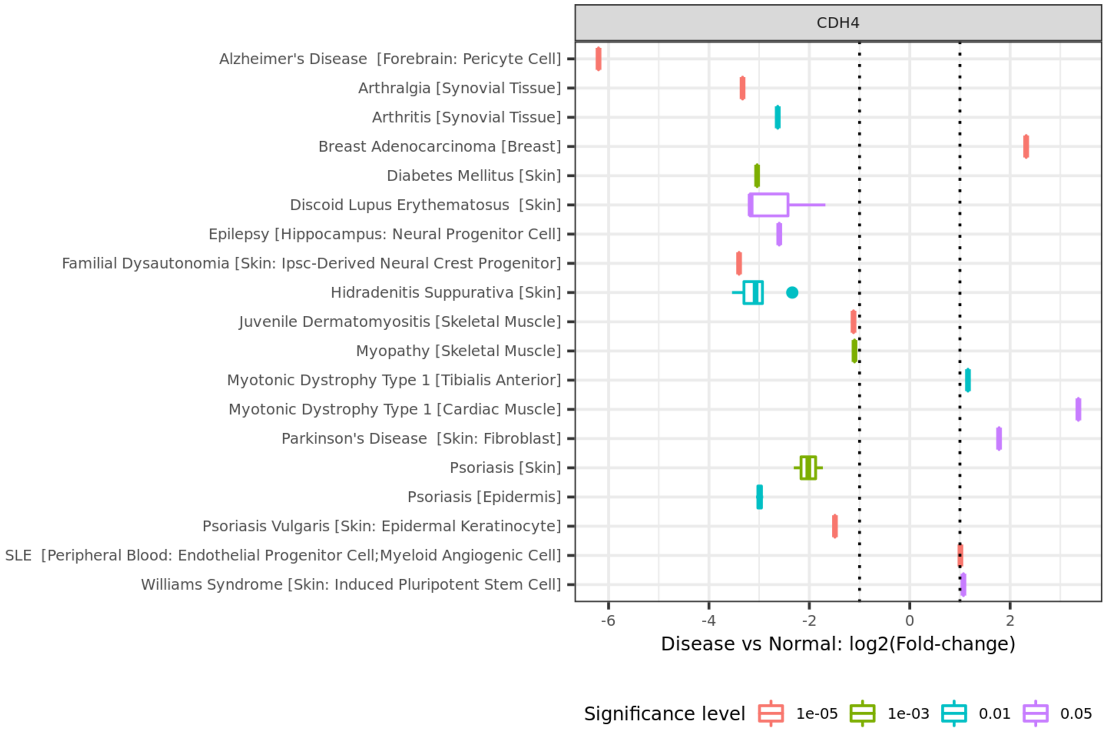

CDH4
Alzheimer's Disease [Forebrain: Pericyte Cell]
Arthralgia [Synovial Tissue]
Arthritis [Synovial Tissue]
Breast Adenocarcinoma [Breast]
Diabetes Mellitus [Skin]
Discoid Lupus Erythematosus [Skin]
Epilepsy [Hippocampus: Neural Progenitor Cell]
Familial Dysautonomia [Skin: Ipsc-Derived Neural Crest Progenitor]
Hidradenitis Suppurativa [Skin]
Juvenile Dermatomyositis [Skeletal Muscle]
Myopathy [Skeletal Muscle]
Myotonic Dystrophy Type 1 [Tibialis Anterior]
Myotonic Dystrophy Type 1 [Cardiac Muscle]
Parkinson's Disease [Skin: Fibroblast]
Psoriasis [Skin]
Psoriasis [Epidermis]
Psoriasis Vulgaris [Skin: Epidermal Keratinocyte]
SLE [Peripheral Blood: Endothelial Progenitor Cell;Myeloid Angiogenic Cell]
Williams Syndrome [Skin: Induced Pluripotent Stem Cell]
-6
-4
-2
0
2
Disease vs Normal: log2(Fold-change)
Significance level
1e-05
1e-03
0.01
0.05


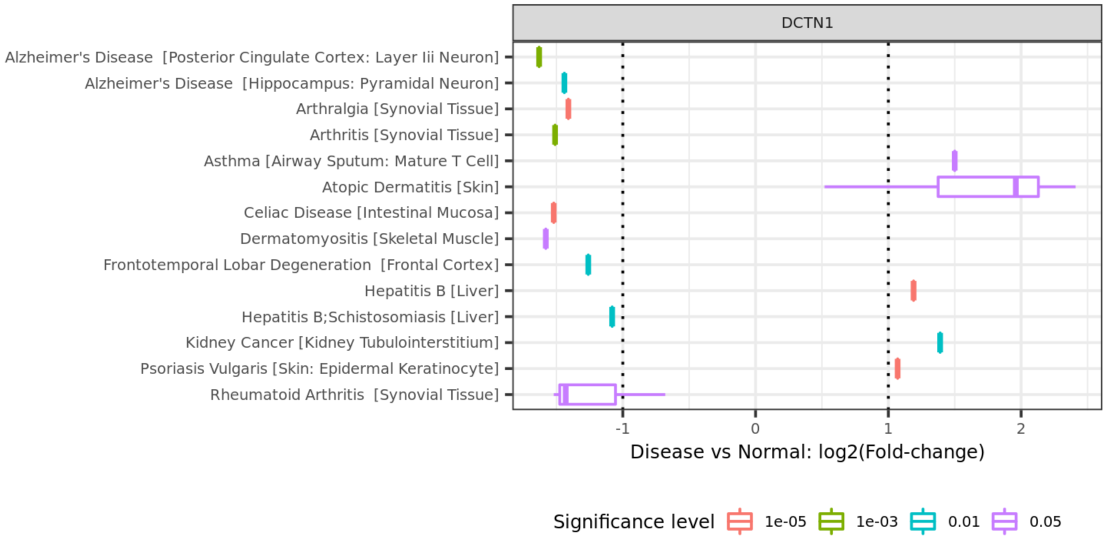

DCTN1
Alzheimer's Disease [Posterior Cingulate Cortex: Layer Iii Neuron]
Alzheimer's Disease [Hippocampus: Pyramidal Neuron]
Arthralgia [Synovial Tissue]
Arthritis [Synovial Tissue]
Asthma [Airway Sputum: Mature T Cell]
Atopic Dermatitis [Skin]
Celiac Disease [Intestinal Mucosa]
Dermatomyositis [Skeletal Muscle]
Frontotemporal Lobar Degeneration [Frontal Cortex]
Hepatitis B [Liver]
Hepatitis B;Schistosomiasis [Liver]
Kidney Cancer [Kidney Tubulointerstitium]
Psoriasis Vulgaris [Skin: Epidermal Keratinocyte]
Rheumatoid Arthritis [Synovial Tissue]
-1
0
1
2
Disease vs Normal: log2(Fold-change)
Significance level
1e-05
1e-03
0.01
0.05

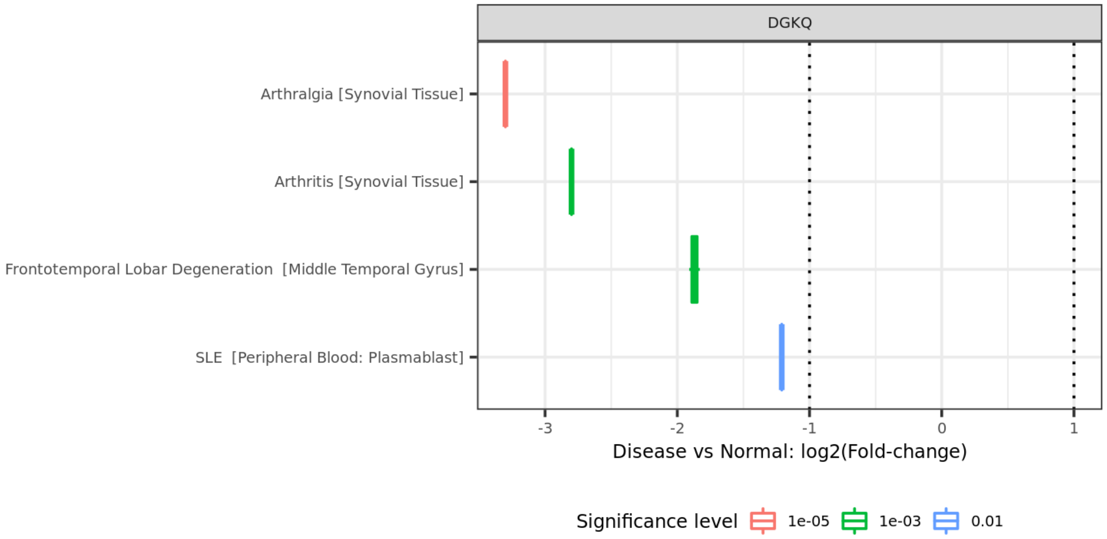
DGKQ
Arthralgia [Synovial Tissue]
Arthritis [Synovial Tissue]
Frontotemporal Lobar Degeneration [Middle Temporal Gyrus]
SLE [Peripheral Blood: Plasmablast]
-3
-2
-1
0
1
Disease vs Normal: log2(Fold-change)
Significance level
1e-05
1e-03
0.01

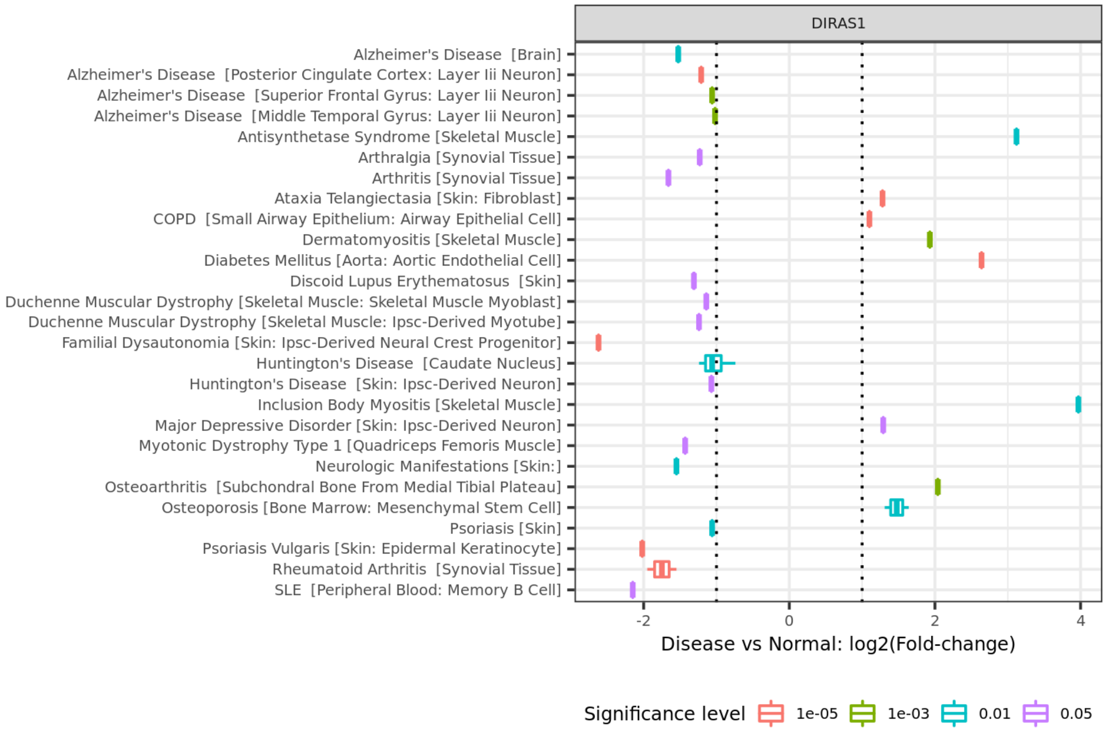
DIRAS1
Alzheimer's Disease [Brain]
Alzheimer's Disease [Posterior Cingulate Cortex: Layer Iii Neuron]
Alzheimer's Disease [Superior Frontal Gyrus: Layer Iii Neuron]
Alzheimer's Disease [Middle Temporal Gyrus: Layer Iii Neuron]
Antisynthetase Syndrome [Skeletal Muscle]
Arthralgia [Synovial Tissue]
Arthritis [Synovial Tissue]
Ataxia Telangiectasia [Skin: Fibroblast]
COPD [Small Airway Epithelium: Airway Epithelial Cell]
Dermatomyositis [Skeletal Muscle]
Diabetes Mellitus [Aorta: Aortic Endothelial Cell]
Discoid Lupus Erythematosus [Skin]
Duchenne Muscular Dystrophy [Skeletal Muscle: Skeletal Muscle Myoblast]
Duchenne Muscular Dystrophy [Skeletal Muscle: Ipsc-Derived Myotube]
Familial Dysautonomia [Skin: Ipsc-Derived Neural Crest Progenitor]
Huntington's Disease [Caudate Nucleus]
Huntington's Disease [Skin: Ipsc-Derived Neuron]
Inclusion Body Myositis [Skeletal Muscle]
Major Depressive Disorder [Skin: Ipsc-Derived Neuron]
Myotonic Dystrophy Type 1 [Quadriceps Femoris Muscle]
Neurologic Manifestations [Skin:]
Osteoarthritis [Subchondral Bone From Medial Tibial Plateau]
Osteoporosis [Bone Marrow: Mesenchymal Stem Cell]
Psoriasis [Skin]
Psoriasis Vulgaris [Skin: Epidermal Keratinocyte]
Rheumatoid Arthritis [Synovial Tissue]
SLE [Peripheral Blood: Memory B Cell]
-2
0
2
4
Disease vs Normal: log2(Fold-change)
Significance level
1e-05
1e-03
0.01
0.05

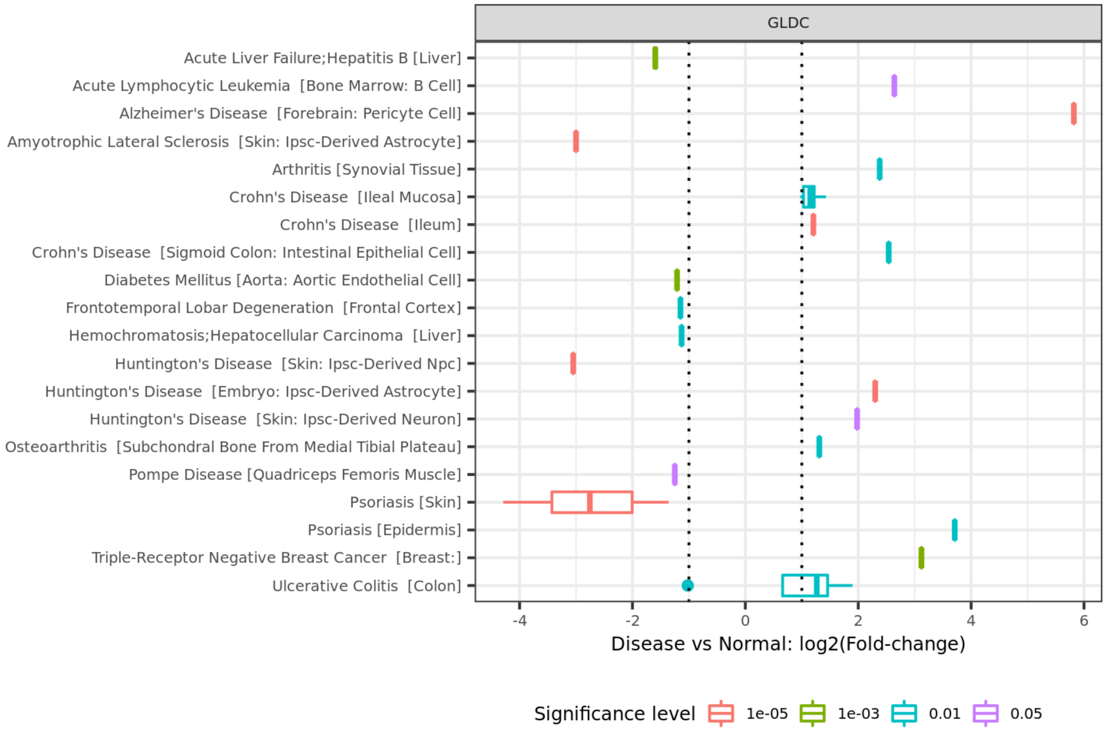
GLDC
Acute Liver Failure;Hepatitis B [Liver]
Acute Lymphocytic Leukemia [Bone Marrow: B Cell]
Alzheimer's Disease [Forebrain: Pericyte Cell]
Amyotrophic Lateral Sclerosis [Skin: Ipsc-Derived Astrocyte]
Arthritis [Synovial Tissue]
Crohn's Disease [Ileal Mucosa]
Crohn's Disease [Ileum]
Crohn's Disease [Sigmoid Colon: Intestinal Epithelial Cell]
Diabetes Mellitus [Aorta: Aortic Endothelial Cell]
Frontotemporal Lobar Degeneration [Frontal Cortex]
Hemochromatosis;Hepatocellular Carcinoma [Liver]
Huntington's Disease [Skin: Ipsc-Derived Npc]
Huntington's Disease [Embryo: Ipsc-Derived Astrocyte]
Huntington's Disease [Skin: Ipsc-Derived Neuron]
Osteoarthritis [Subchondral Bone From Medial Tibial Plateau]
Pompe Disease [Quadriceps Femoris Muscle]
Psoriasis [Skin]
Psoriasis [Epidermis]
Triple-Receptor Negative Breast Cancer [Breast:]
Ulcerative Colitis [Colon]
-4
-2
0
2
4
6
Disease vs Normal: log2(Fold-change)
Significance level
1e-05
1e-03
0.01
0.05

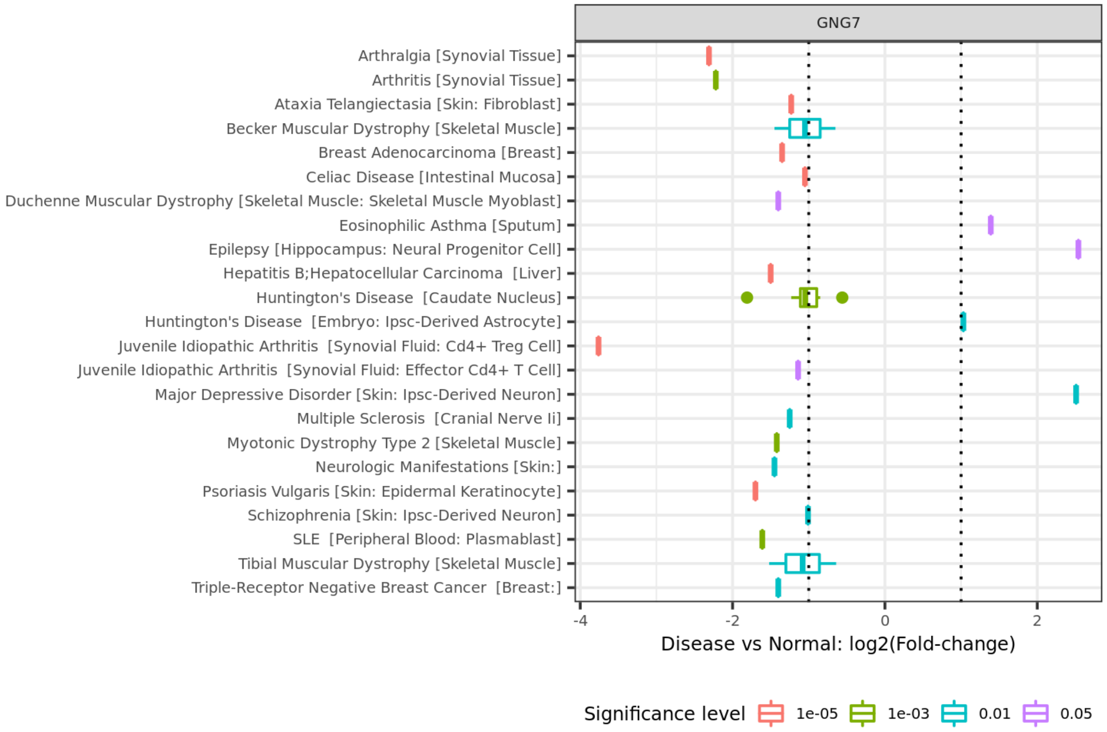
GNG7
Arthralgia [Synovial Tissue]
Arthritis [Synovial Tissue]
Ataxia Telangiectasia [Skin: Fibroblast]
Becker Muscular Dystrophy [Skeletal Muscle]
Breast Adenocarcinoma [Breast]
Celiac Disease [Intestinal Mucosa]
Duchenne Muscular Dystrophy [Skeletal Muscle: Skeletal Muscle Myoblast]
Eosinophilic Asthma [Sputum]
Epilepsy [Hippocampus: Neural Progenitor Cell]
Hepatitis B;Hepatocellular Carcinoma [Liver]
Huntington's Disease [Caudate Nucleus]
Huntington's Disease [Embryo: Ipsc-Derived Astrocyte]
Juvenile Idiopathic Arthritis [Synovial Fluid: Cd4+ Treg Cell]
Juvenile Idiopathic Arthritis [Synovial Fluid: Effector Cd4+ T Cell]
Major Depressive Disorder [Skin: Ipsc-Derived Neuron]
Multiple Sclerosis [Cranial Nerve Ii]
Myotonic Dystrophy Type 2 [Skeletal Muscle]
Neurologic Manifestations [Skin:]
Psoriasis Vulgaris [Skin: Epidermal Keratinocyte]
Schizophrenia [Skin: Ipsc-Derived Neuron]
SLE [Peripheral Blood: Plasmablast]
Tibial Muscular Dystrophy [Skeletal Muscle]
Triple-Receptor Negative Breast Cancer [Breast:]
-4
-2
0
2
Disease vs Normal: log2(Fold-change)
Significance level
1e-05
1e-03
0.01
0.05

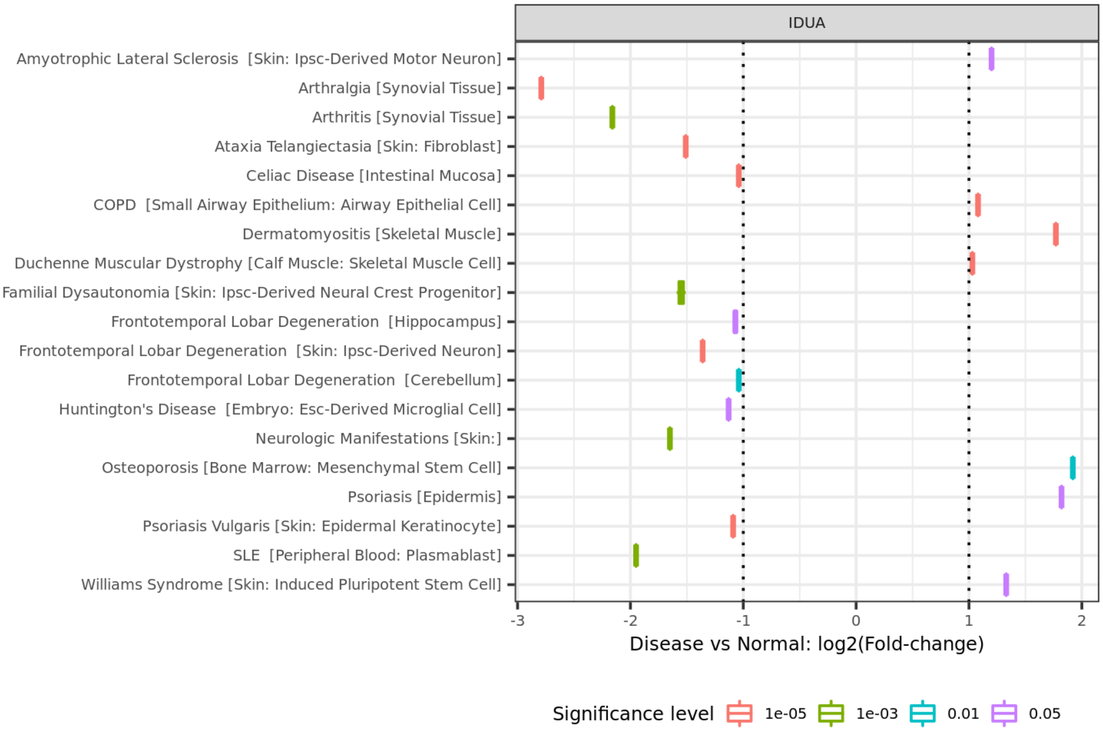
IDUA
Amyotrophic Lateral Sclerosis [Skin: Ipsc-Derived Motor Neuron]
Arthralgia [Synovial Tissue]
Arthritis [Synovial Tissue]
Ataxia Telangiectasia [Skin: Fibroblast]
Celiac Disease [Intestinal Mucosa]
COPD [Small Airway Epithelium: Airway Epithelial Cell]
Dermatomyositis [Skeletal Muscle]
Duchenne Muscular Dystrophy [Calf Muscle: Skeletal Muscle Cell]
Familial Dysautonomia [Skin: Ipsc-Derived Neural Crest Progenitor]
Frontotemporal Lobar Degeneration [Hippocampus]
Frontotemporal Lobar Degeneration [Skin: Ipsc-Derived Neuron]
Frontotemporal Lobar Degeneration [Cerebellum]
Huntington's Disease [Embryo: Esc-Derived Microglial Cell]
Neurologic Manifestations [Skin:]
Osteoporosis [Bone Marrow: Mesenchymal Stem Cell]
Psoriasis [Epidermis]
Psoriasis Vulgaris [Skin: Epidermal Keratinocyte]
SLE [Peripheral Blood: Plasmablast]
Williams Syndrome [Skin: Induced Pluripotent Stem Cell]
-3
-2
-1
0
1
2
Disease vs Normal: log2(Fold-change)
Significance level
1e-05
1e-03
0.01
0.05

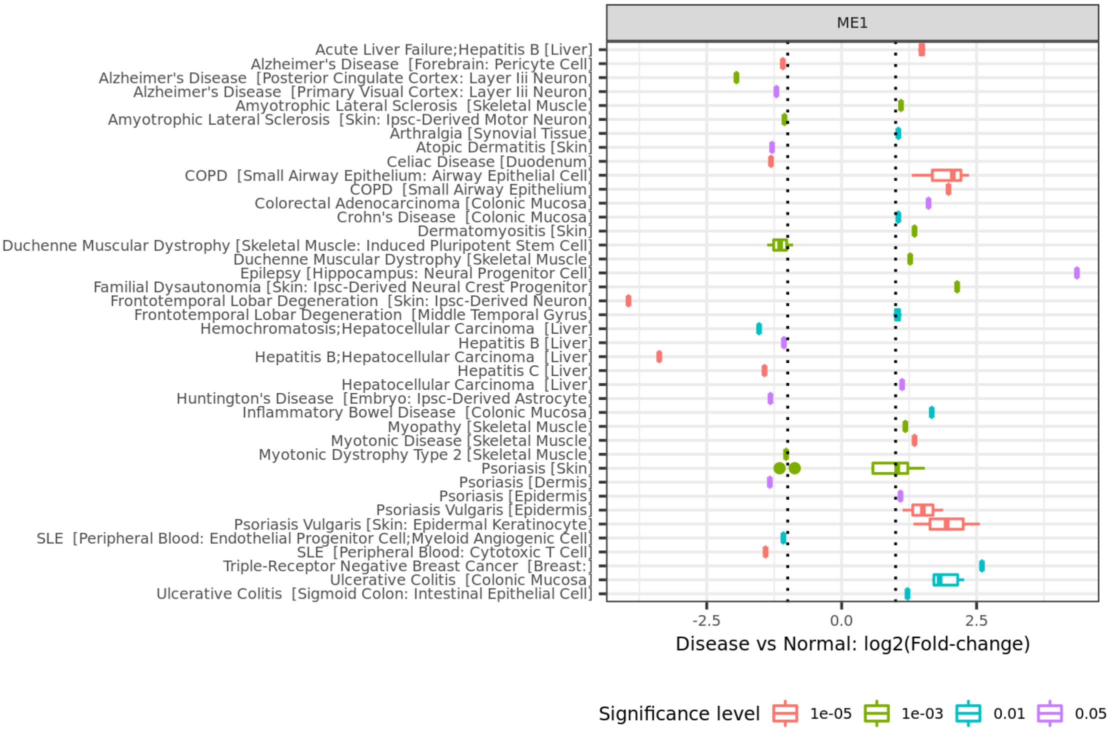
ME1
Acute Liver Failure;Hepatitis B [Liver]
Alzheimer's Disease [Forebrain: Pericyte Cell]
Alzheimer's Disease [Posterior Cingulate Cortex: Layer Iii Neuron]
Alzheimer's Disease [Primary Visual Cortex: Layer Iii Neuron]
Amyotrophic Lateral Sclerosis [Skeletal Muscle]
Amyotrophic Lateral Sclerosis [Skin: Ipsc-Derived Motor Neuron]
Arthralgia [Synovial Tissue]
Atopic Dermatitis [Skin]
Celiac Disease [Duodenum]
COPD [Small Airway Epithelium: Airway Epithelial Cell]
COPD [Small Airway Epithelium]
Colorectal Adenocarcinoma [Colonic Mucosa]
Crohn's Disease [Colonic Mucosa]
Dermatomyositis [Skin]
Duchenne Muscular Dystrophy [Skeletal Muscle: Induced Pluripotent Stem Cell]
Duchenne Muscular Dystrophy [Skeletal Muscle]
Epilepsy [Hippocampus: Neural Progenitor Cell]
Familial Dysautonomia [Skin: Ipsc-Derived Neural Crest Progenitor]
Frontotemporal Lobar Degeneration [Skin: Ipsc-Derived Neuron]
Frontotemporal Lobar Degeneration [Middle Temporal Gyrus]
Hemochromatosis;Hepatocellular Carcinoma [Liver]
Hepatitis B [Liver]
Hepatitis B;Hepatocellular Carcinoma [Liver]
Hepatitis C [Liver]
Hepatocellular Carcinoma [Liver]
Huntington's Disease [Embryo: Ipsc-Derived Astrocyte]
Inflammatory Bowel Disease [Colonic Mucosa]
Myopathy [Skeletal Muscle]
Myotonic Disease [Skeletal Muscle]
Myotonic Dystrophy Type 2 [Skeletal Muscle]
Psoriasis [Skin]
Psoriasis [Dermis]
Psoriasis [Epidermis]
Psoriasis Vulgaris [Epidermis]
Psoriasis Vulgaris [Skin: Epidermal Keratinocyte]
SLE [Peripheral Blood: Endothelial Progenitor Cell;Myeloid Angiogenic Cell]
SLE [Peripheral Blood: Cytotoxic T Cell]
Triple-Receptor Negative Breast Cancer [Breast:]
Ulcerative Colitis [Colonic Mucosa]
Ulcerative Colitis [Sigmoid Colon: Intestinal Epithelial Cell]
-2.5
0.0
2.5
Disease vs Normal: log2(Fold-change)
Significance level
1e-05
1e-03
0.01
0.05

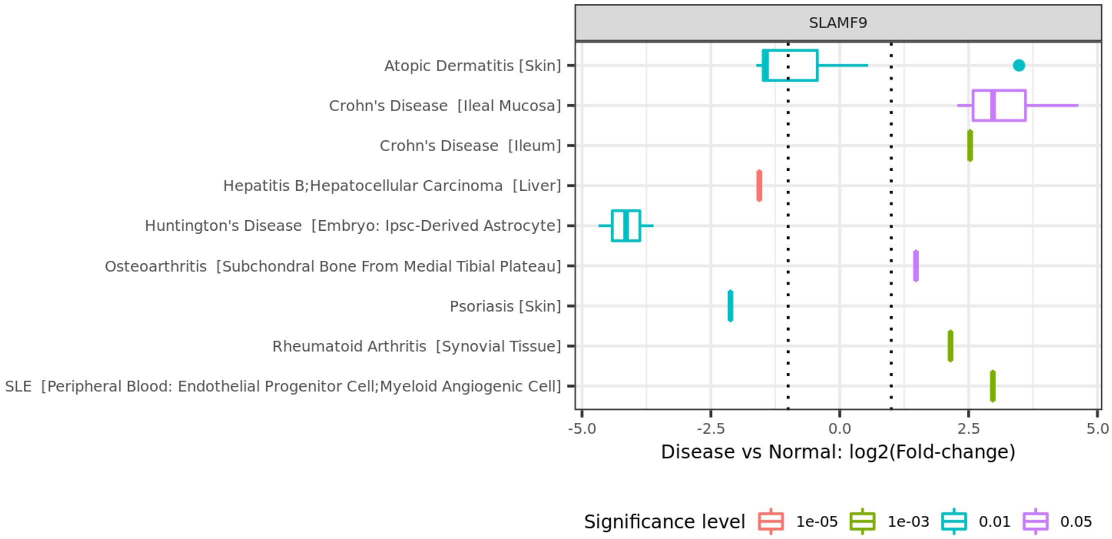
SLAMF9
Atopic Dermatitis [Skin]
Crohn's Disease [Ileal Mucosa]
Crohn's Disease [Ileum]
Hepatitis B;Hepatocellular Carcinoma [Liver]
Huntington's Disease [Embryo: Ipsc-Derived Astrocyte]
Osteoarthritis [Subchondral Bone From Medial Tibial Plateau]
Psoriasis [Skin]
Rheumatoid Arthritis [Synovial Tissue]
SLE [Peripheral Blood: Endothelial Progenitor Cell;Myeloid Angiogenic Cell]
-5.0
-2.5
0.0
2.5
5.0
Disease vs Normal: log2(Fold-change)
Significance level
1e-05
1e-03
0.01
0.05

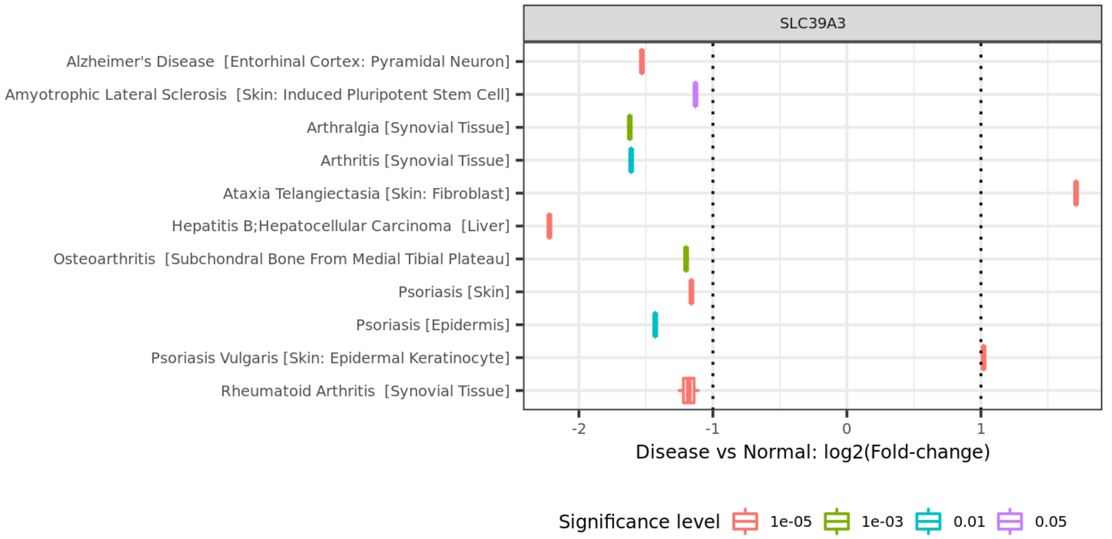
SLC39A3
Alzheimer's Disease [Entorhinal Cortex: Pyramidal Neuron]
Amyotrophic Lateral Sclerosis [Skin: Induced Pluripotent Stem Cell]
Arthralgia [Synovial Tissue]
Arthritis [Synovial Tissue]
Ataxia Telangiectasia [Skin: Fibroblast]
Hepatitis B;Hepatocellular Carcinoma [Liver]
Osteoarthritis [Subchondral Bone From Medial Tibial Plateau]
Psoriasis [Skin]
Psoriasis [Epidermis]
Psoriasis Vulgaris [Skin: Epidermal Keratinocyte]
Rheumatoid Arthritis [Synovial Tissue]
-2
-1
0
1
Disease vs Normal: log2(Fold-change)
Significance level
1e-05
1e-03
0.01
0.05

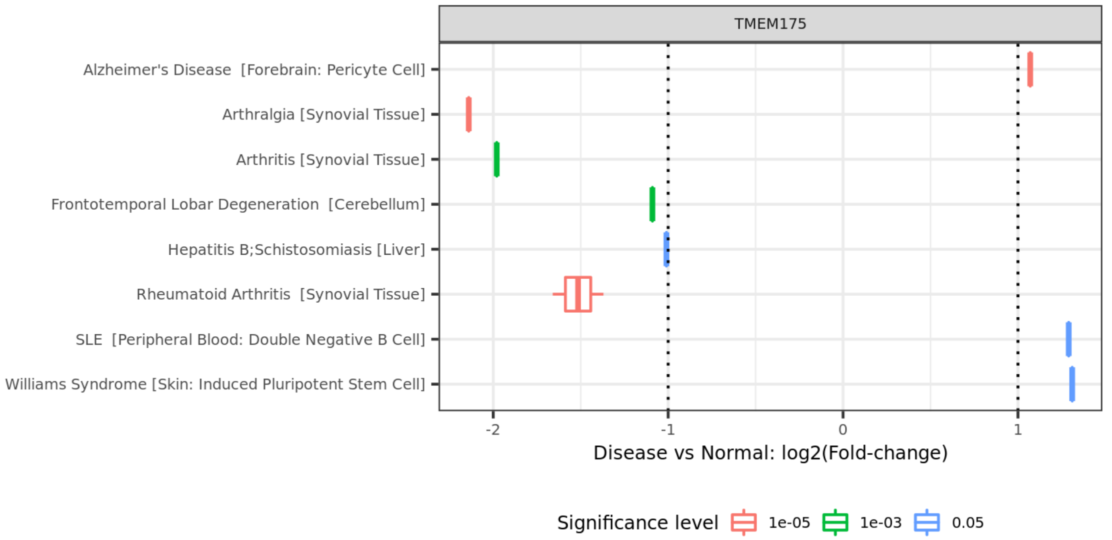
TMEM175
Alzheimer's Disease [Forebrain: Pericyte Cell]
Arthralgia [Synovial Tissue]
Arthritis [Synovial Tissue]
Frontotemporal Lobar Degeneration [Cerebellum]
Hepatitis B;Schistosomiasis [Liver]
Rheumatoid Arthritis [Synovial Tissue]
SLE [Peripheral Blood: Double Negative B Cell]
Williams Syndrome [Skin: Induced Pluripotent Stem Cell]
-2
-1
0
1
Disease vs Normal: log2(Fold-change)
Significance level
1e-05
1e-03
0.05

Supplementary figure 3. Differential gene expression results for CIDP or IP related genes identified in MR and Colocalization analyses.
