## Supplementary figure 4-5 for "Genetic analyses of inflammatory polyneuropathy and chronic inflammatory demyelinating polyradiculoneuropathy identified candidate genes"

Bulk tissue gene expression for CDH4 (ENSG00000179242.15)

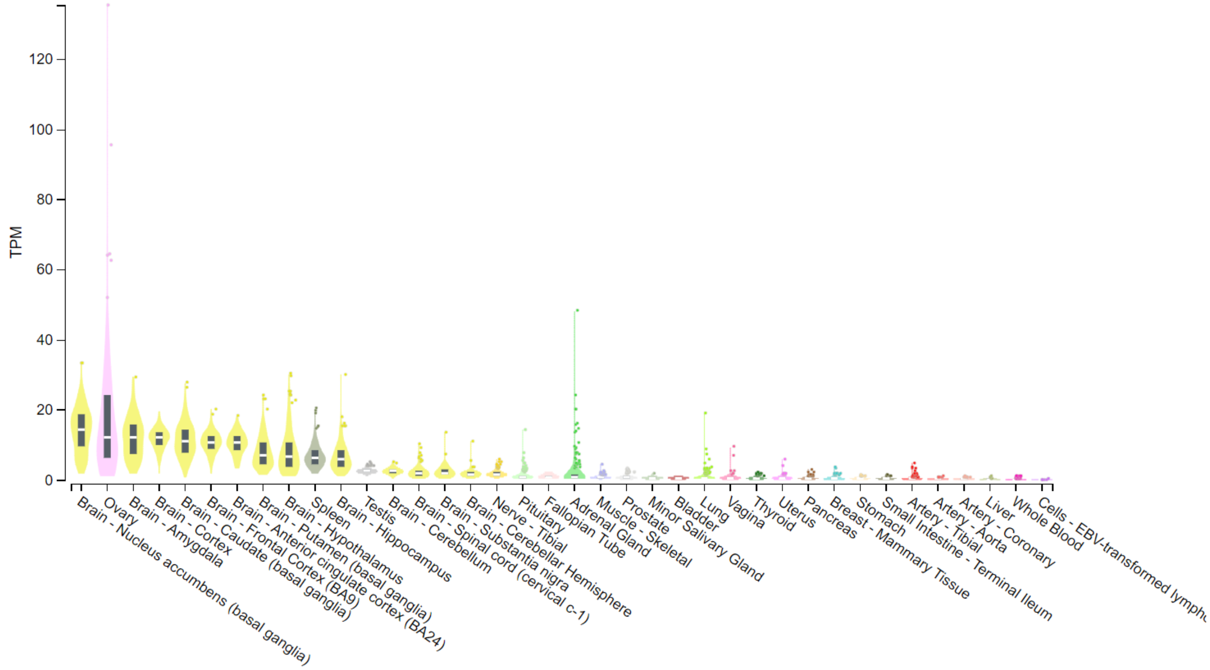


Bulk tissue gene expression for DGKQ (ENSG00000145214.13)

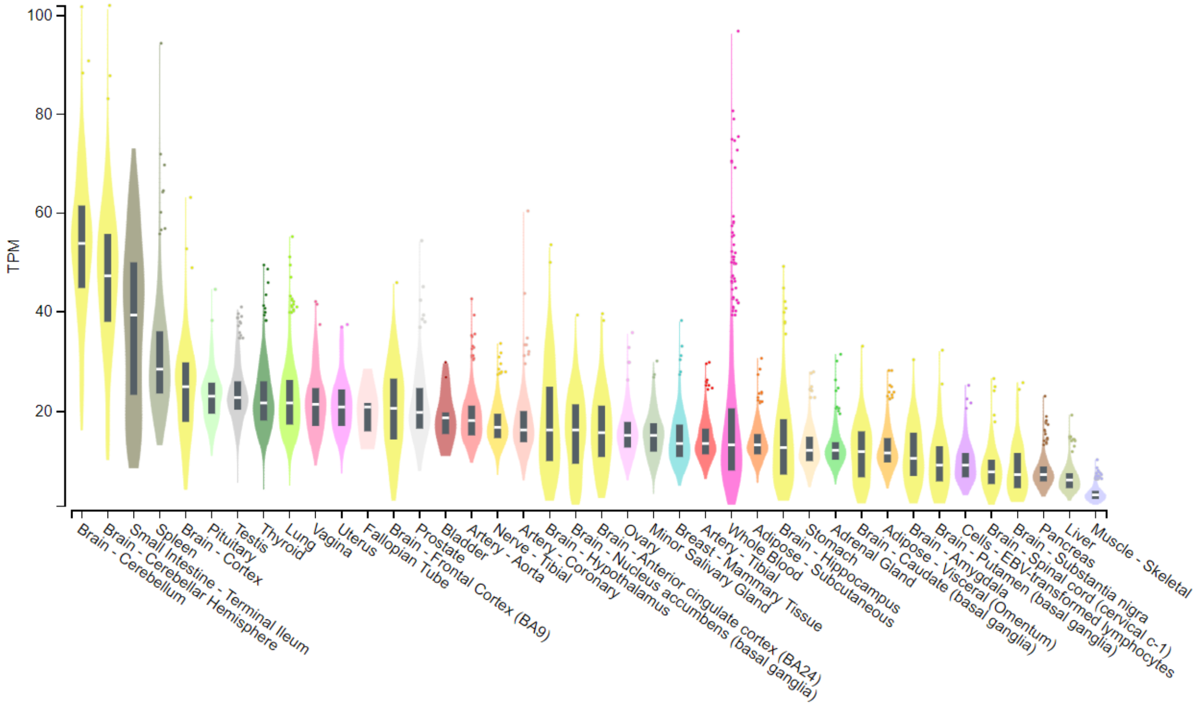

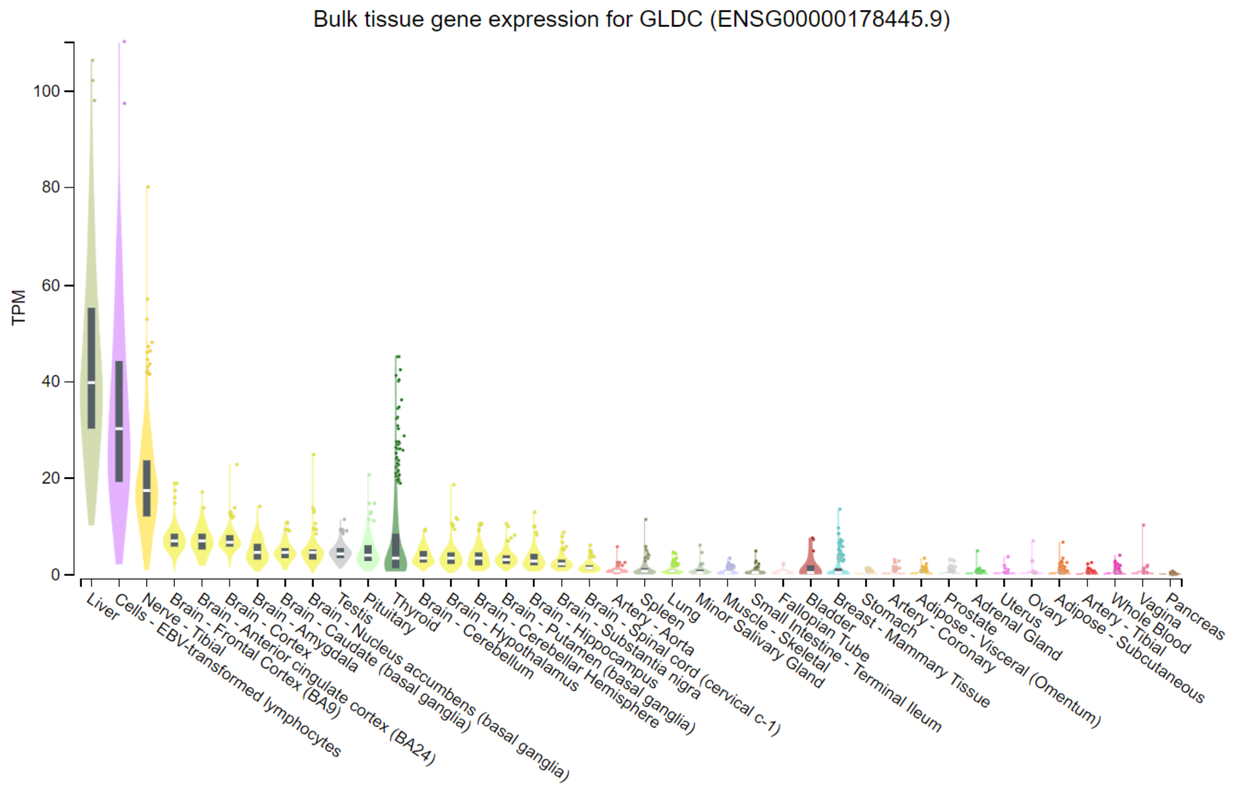
Bulk tissue gene expression for GLDC (ENSG00000178445.9)
TPM
0
20
40
60
80
100
Liver
Cells - EBV-transformed lymphocytes
Nerve - Tibial
Brain - Frontal Cortex (BA9)
Brain - Anterior cingulate cortex (BA24)
Brain - Cortex
Brain - Amygdala
Brain - Caudate (basal ganglia)
Brain - Nucleus accumbens (basal ganglia)
Testis
Pituitary
Thyroid
Brain - Cerebellum
Brain - Hypothalamus
Brain - Cerebellar Hemisphere
Brain - Putamen (basal ganglia)
Brain - Hippocampus
Brain - Substantia nigra
Brain - Spinal cord (cervical c-1)
Artery - Aorta
Spleen
Lung
Minor Salivary Gland
Muscle - Skeletal
Small Intestine - Terminal Ileum
Fallopian Tube
Bladder
Breast - Mammary Tissue
Stomach
Artery - Coronary
Adipose - Visceral (Omentum)
Prostate
Adrenal Gland
Uterus
Ovary
Adipose - Subcutaneous
Artery - Tibial
Whole Blood
Vagina
Pancreas

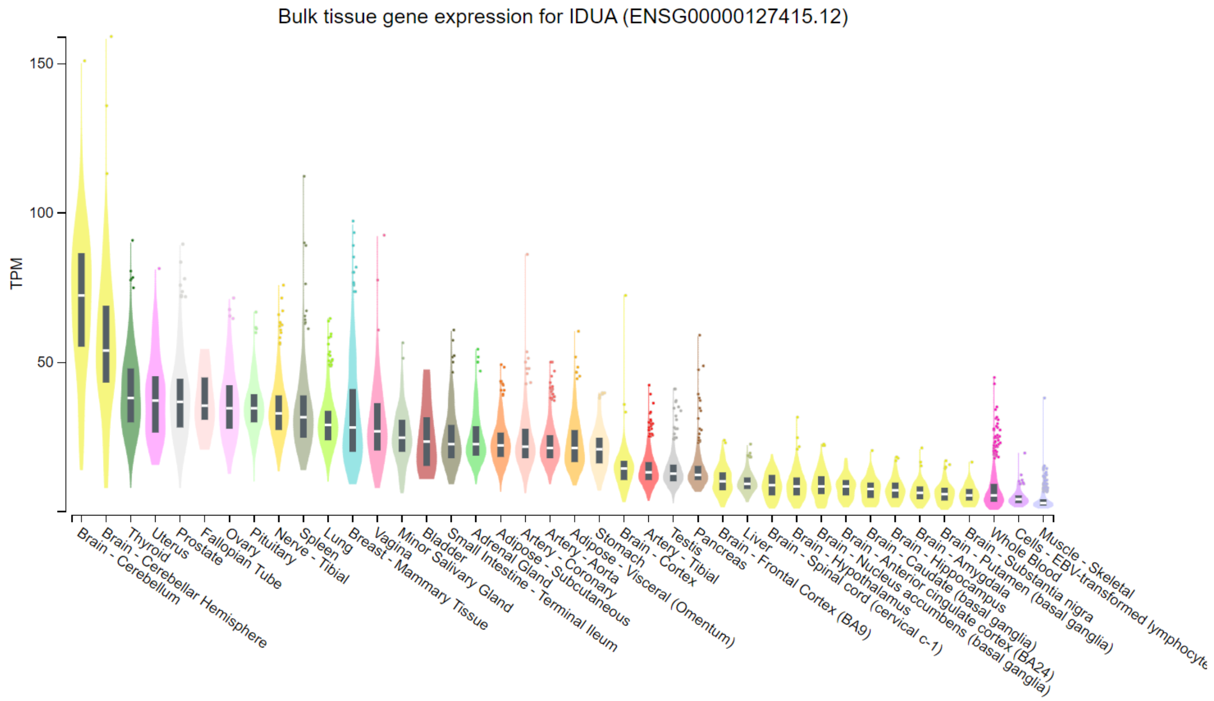
Bulk tissue gene expression for IDUA (ENSG00000127415.12)
TPM
50
100
150
Brain - Cerebellum
Brain - Cerebellar Hemisphere
Thyroid
Uterus
Prostate
Fallopian Tube
Ovary
Pituitary
Nerve - Tibial
Spleen
Lung
Breast - Mammary Tissue
Vagina
Minor Salivary Gland
Bladder
Small Intestine - Terminal Ileum
Adrenal Gland
Adipose - Subcutaneous
Artery - Coronary
Artery - Aorta
Adipose - Visceral (Omentum)
Stomach
Brain - Cortex
Artery - Tibial
Testis
Pancreas
Brain - Frontal Cortex (BA9)
Liver
Brain - Spinal cord (cervical c-1)
Brain - Hypothalamus
Brain - Nucleus accumbens (basal ganglia)
Brain - Anterior cingulate cortex (BA24)
Brain - Caudate (basal ganglia)
Brain - Hippocampus
Brain - Amygdala
Brain - Putamen (basal ganglia)
Brain - Substantia nigra
Whole Blood
Cells - EBV-transformed lymphocyte
Muscle - Skeletal

Bulk tissue gene expression for SLAMF8 (ENSG00000158714.10)

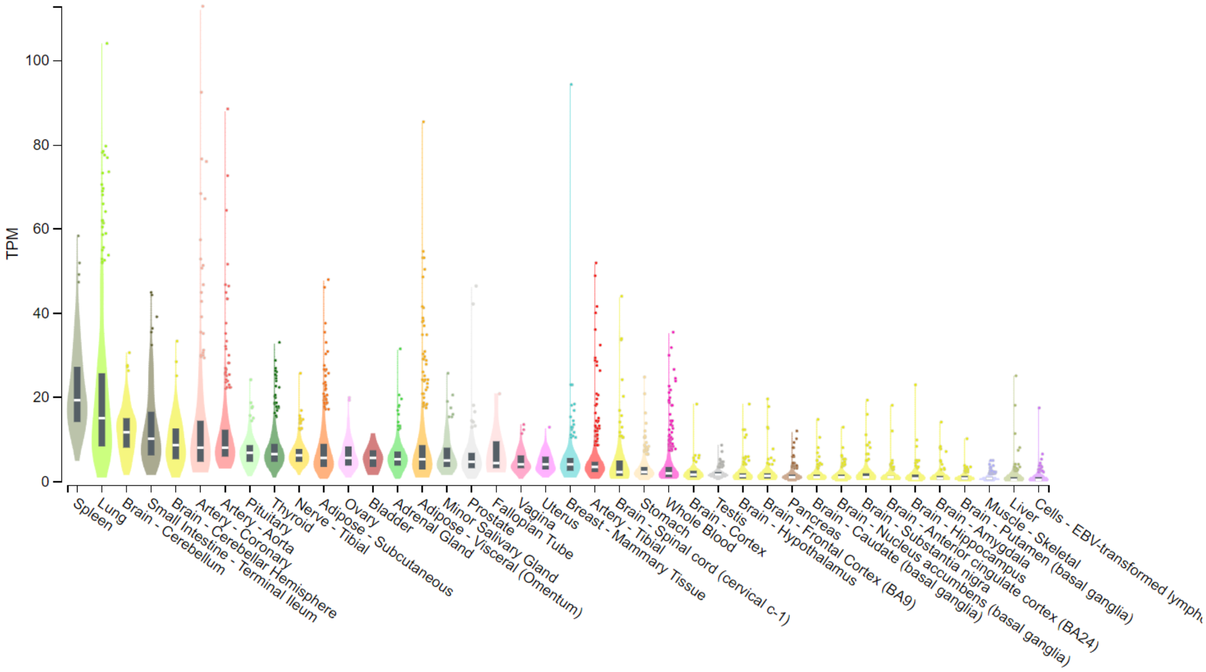


Bulk tissue gene expression for TMEM175 (ENSG00000127419.16)

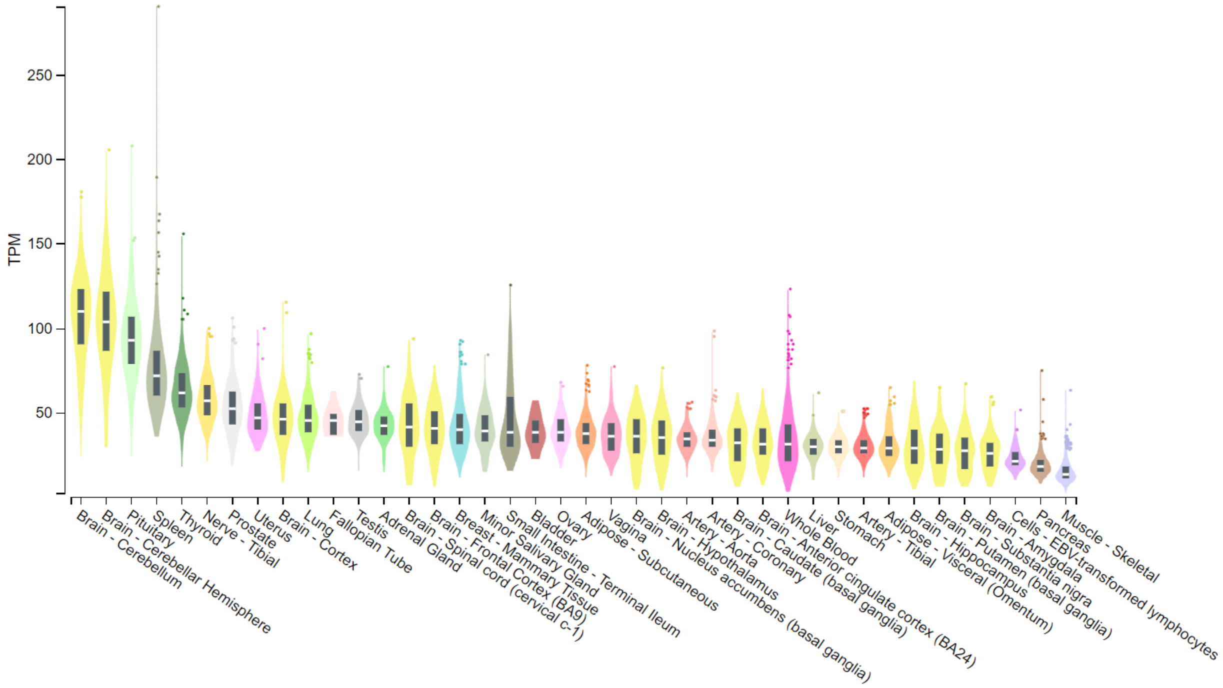

Bulk tissue gene expression for DIRAS1 (ENSG00000176490.4)

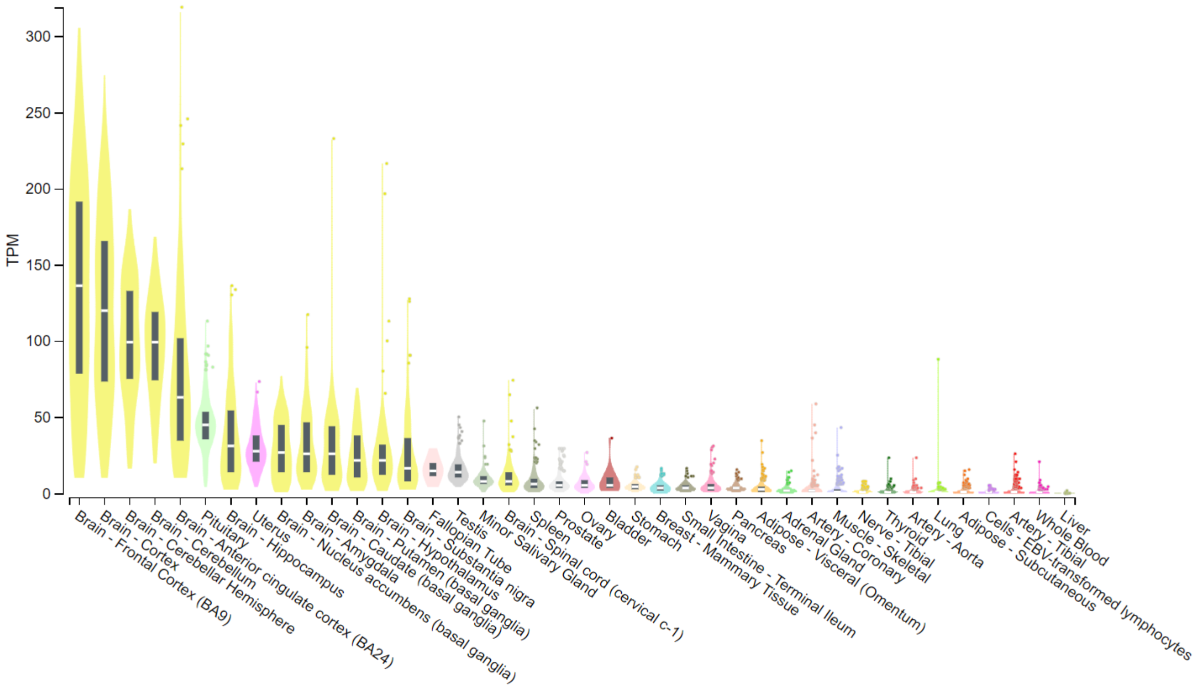


Bulk tissue gene expression for GNG7 (ENSG00000176533.12)

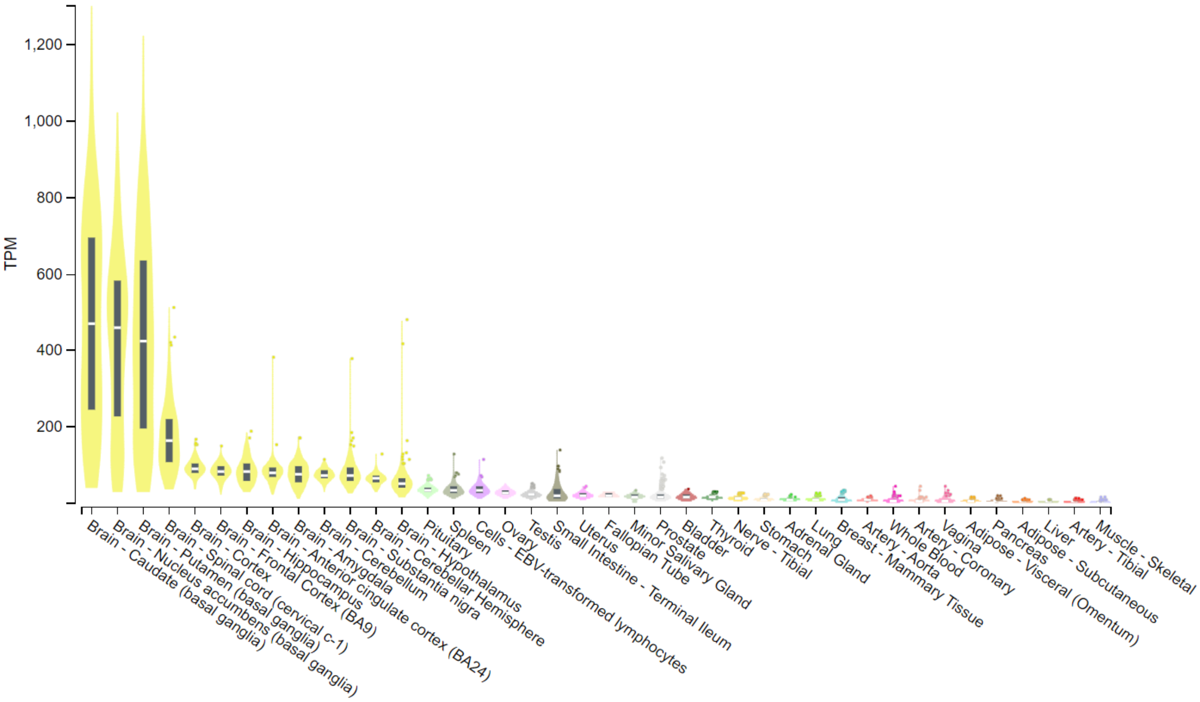

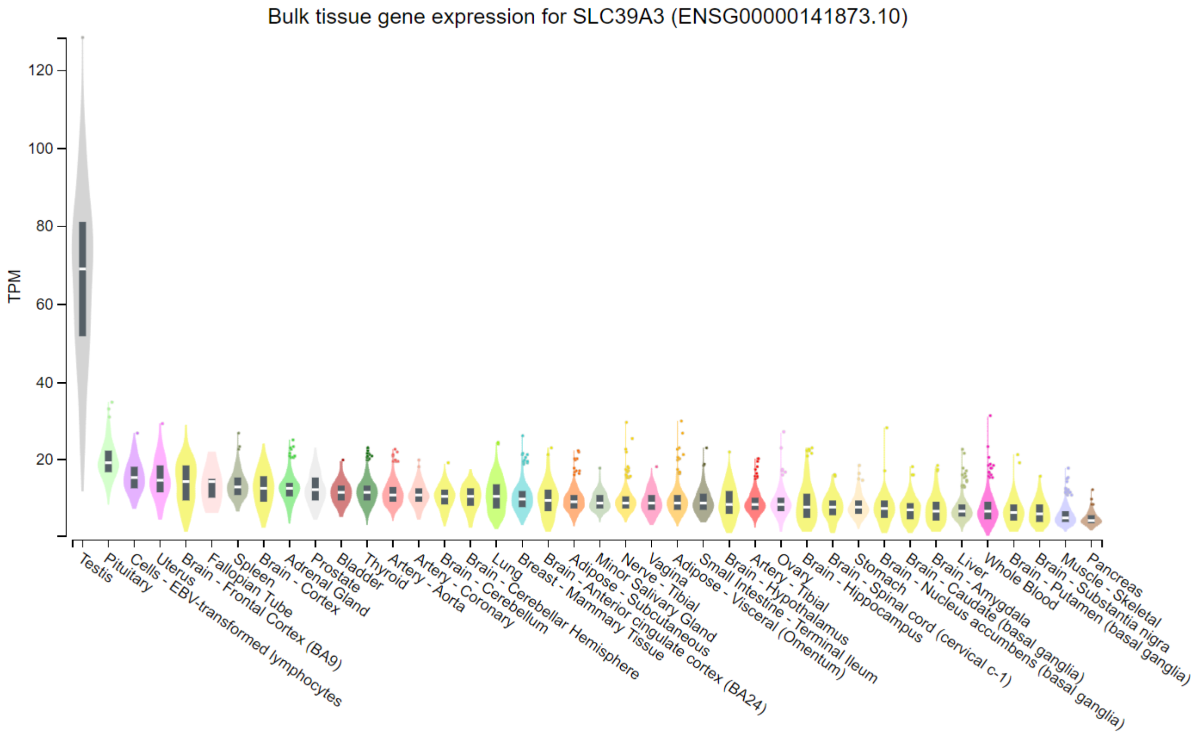
Bulk tissue gene expression for SLC39A3 (ENSG00000141873.10)
TPM
120
100
80
60
40
20
Testis
Pituitary
Cells - EBV-transformed lymphocytes
Uterus
Brain - Frontal Cortex (BA9)
Fallopian Tube
Spleen
Brain - Cortex
Adrenal Gland
Prostate
Bladder
Thyroid
Artery - Aorta
Artery - Coronary
Brain - Cerebellum
Lung
Brain - Cerebellar Hemisphere
Breast - Mammary Tissue
Brain - Anterior cingulate cortex (BA24)
Adipose - Subcutaneous
Minor Salivary Gland
Nerve - Tibial
Vagina
Adipose - Visceral (Omentum)
Small Intestine - Terminal Ileum
Brain - Hypothalamus
Artery - Tibial
Ovary
Brain - Hippocampus
Stomach
Brain - Spinal cord (cervical c-1)
Brain - Nucleus accumbens (basal ganglia)
Brain - Caudate (basal ganglia)
Brain - Amygdala
Liver
Whole Blood
Brain - Putamen (basal ganglia)
Brain - Substantia nigra
Muscle - Skeletal
Pancreas

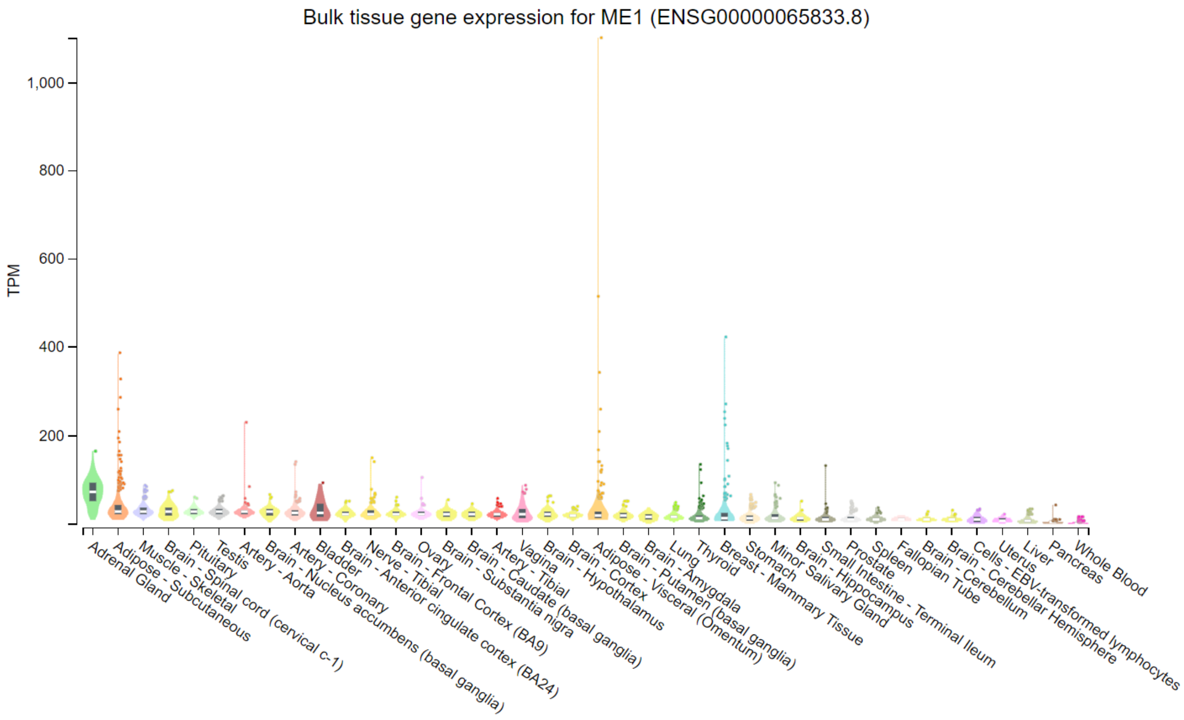
Bulk tissue gene expression for ME1 (ENSG00000065833.8)
TPM
1,000
800
600
400
200
Adrenal Gland
Adipose - Subcutaneous
Muscle - Skeletal
Brain - Spinal cord (cervical c-1)
Pituitary
Testis
Artery - Aorta
Brain - Nucleus accumbens (basal ganglia)
Artery - Coronary
Bladder
Brain - Anterior cingulate cortex (BA24)
Nerve - Tibial
Ovary
Brain - Frontal Cortex (BA9)
Brain - Substantia nigra
Artery - Tibial
Brain - Caudate (basal ganglia)
Vagina
Brain - Hypothalamus
Brain - Cortex
Adipose - Visceral (Omentum)
Brain - Putamen (basal ganglia)
Brain - Amygdala
Lung
Thyroid
Breast - Mammary Tissue
Stomach
Minor Salivary Gland
Brain - Hippocampus
Small Intestine - Terminal Ileum
Prostate
Spleen
Fallopian Tube
Brain - Cerebellum
Brain - Cerebellar Hemisphere
Cells - EBV-transformed lymphocytes
Uterus
Liver
Pancreas
Whole Blood

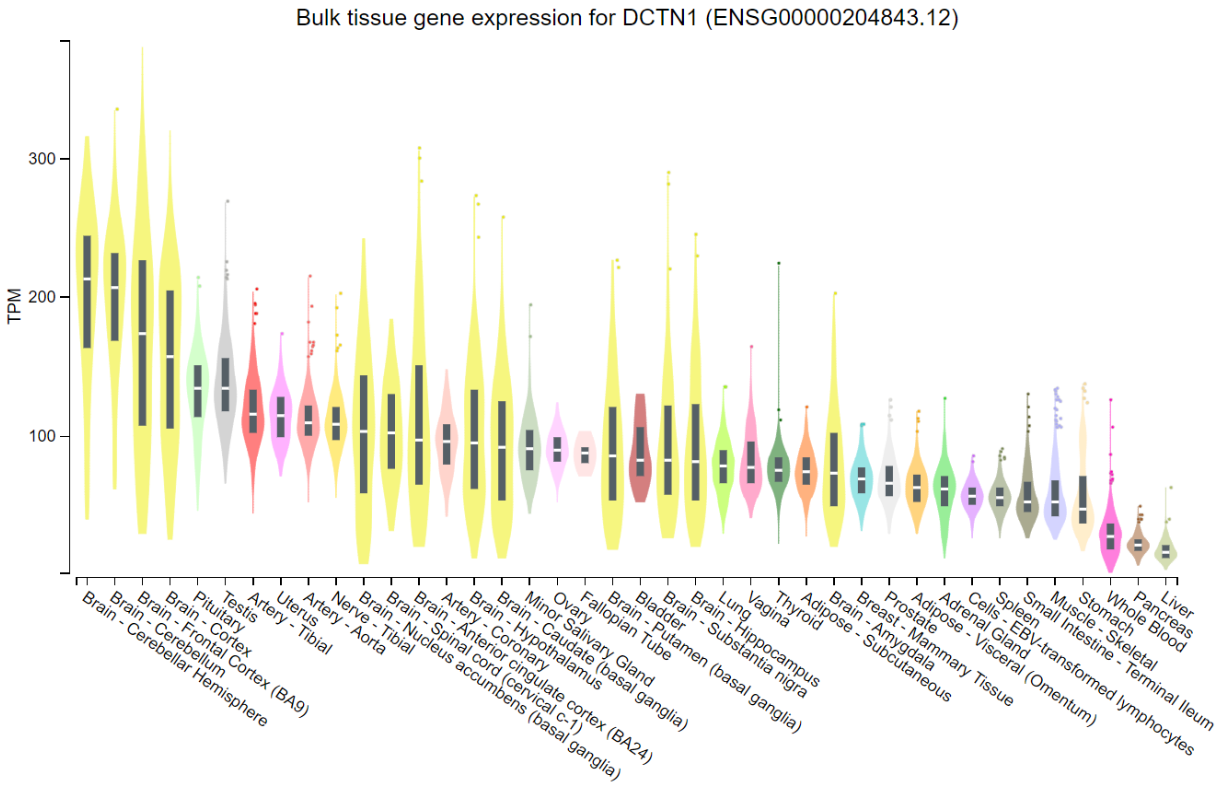


Supplementary figure 4. GTEx bulk tissue gene expression of CIDP/IP -related genes.

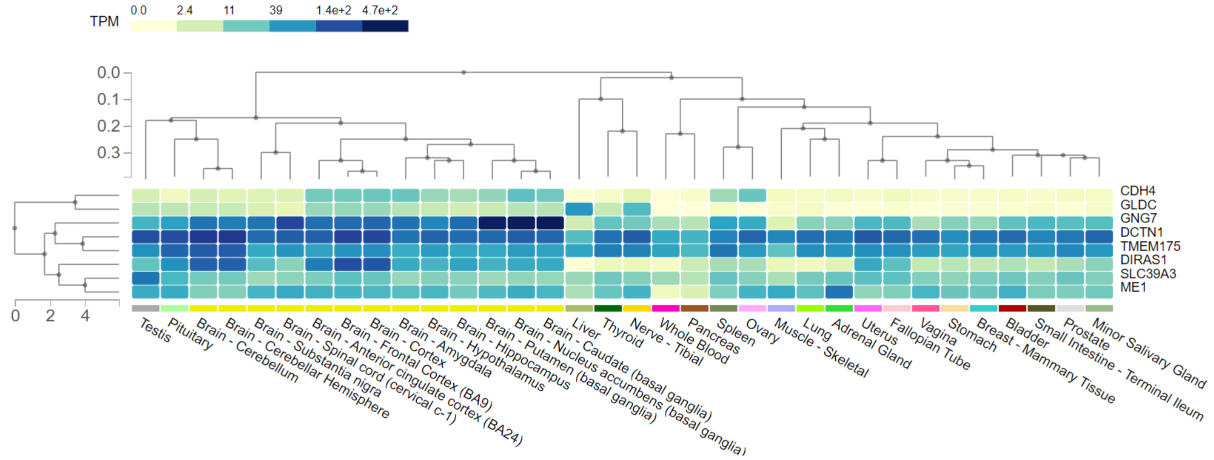


Supplementary figure 5. Heatmap of GTEx bulk tissue gene expression of CIDP/IP -related genes.
